# Implementation of An Online COVID-19 Epidemic Calculator for Tracking the Spread of the Coronavirus in Singapore and Other Countries

**DOI:** 10.1101/2020.06.02.20120188

**Authors:** Fook Fah Yap, Minglee Yong

## Abstract

This paper describes the methods underlying the development of an online COVID-19 Epidemic Calculator for tracking COVID-19 growth parameters. From publicly available infection case data, the calculator is used to estimate the effective reproduction number, doubling time, final epidemic size, and death toll. As a case study, we analyzed the results for Singapore during the “Circuit breaker” period from April 7, 2020 to the end of May 2020. The calculator shows that the stringent measures imposed have an immediate effect of rapidly slowing down the spread of the coronavirus. After about two weeks, the effective reproduction number reduced to 1.0. Since then, the number has been fluctuating around 1.0.

The COVID-19 Epidemic Calculator is available in the form of an online Google Sheet and the results are presented as Tableau Public dashboards at www.cv19.one. By making the calculator readily accessible online, the public can have a tool to meaningfully assess the effectiveness of measures to control the pandemic.

## 1. Imtroduction

As countries around the world take drastic measures to contain the COVID-19 pandemic, people need to understand the effectiveness of such interventions. Making sense of epidemiological data can be challenging given confusing and overlapping terminology. Raw data and statistics on infection numbers (e.g. Figure 1) do not directly help answer the following questions: (1) Are social distancing measures working? (2) How much longer does it take to flatten the curve? (3) What will be the final death toll?

**Figure 1.**
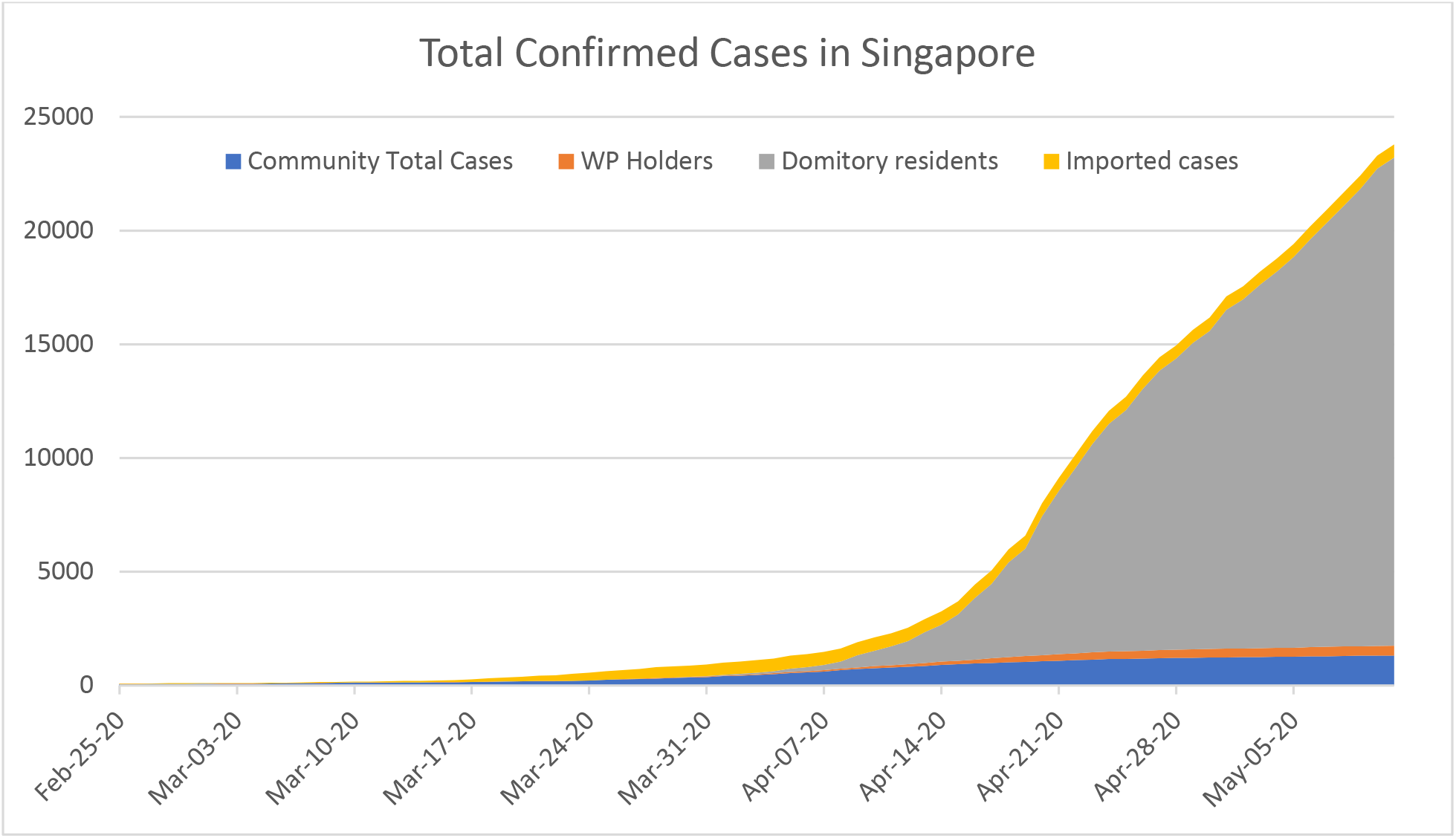
Total Confirmed Cases in Singapore [1].

This paper describes the methods underlying the online COVID-19 Epidemic Calculator for tracking and estimating COVID-19 growth parameters, including reproduction number, doubling time, final epidemic size, and death toll. These methods are illustrated using the case example of Singapore. We demonstrate how the calculator can reveal the effect of imposing strict social distancing measures (“Circuit breaker”) from April 7, 2020 that is not apparent from just looking at infection numbers.

While our methodology is similar to several freely available software packages and programming codes for calculating the effective reproduction number (e.g. [21, 23, 24, 33]), we have implemented the calculations in the widely used Excel spreadsheet. The execution time is a matter of seconds, even for calculating a 4-month, 100-country data set. The calculator is also available as an online Google Sheet to facilitate sharing and collaboration. The input data are obtained from publicly available sources [2, 3, 31] and are updated daily.

The first section of this paper will introduce and define basic terminology for understanding infectious disease transmission given that publicly available data are related to these variables. As the reproduction number is used as an indicator of the effectiveness of interventions, we will demonstrate how to estimate this number using different methods. First, we use the SEIR model while accounting for changing reproduction number and undetected infections. Next, we estimate the effective reproduction number, a more accurate index, using a non-parametric approach and a Bayesian approach. We also show how to derive estimates of dates of actual symptom onset and dates of being exposed which are important for our estimation of the effective reproduction number. To answer the question about the time needed to flatten the curve, we will estimate the doubling time, defined as the time for the total number of infections to double at the current rate. Finally, we describe a method to forecast the final total number of cases and deaths.

## 2. Introduction to terminology

Figure 2 clarifies the different overlapping terminology used in epidemiology and illustrates the timeline for the various stages of infection. *Exposed* is the state at which an individual first becomes infected but is not yet contagious. The *latent period* is the time from being infected (exposed) to becoming contagious. An infected person can be contagious even before the onset of symptoms. Data suggests that some people could have infected others 1 to 3 days before they developed symptoms [10,11].

**Figure 2.**
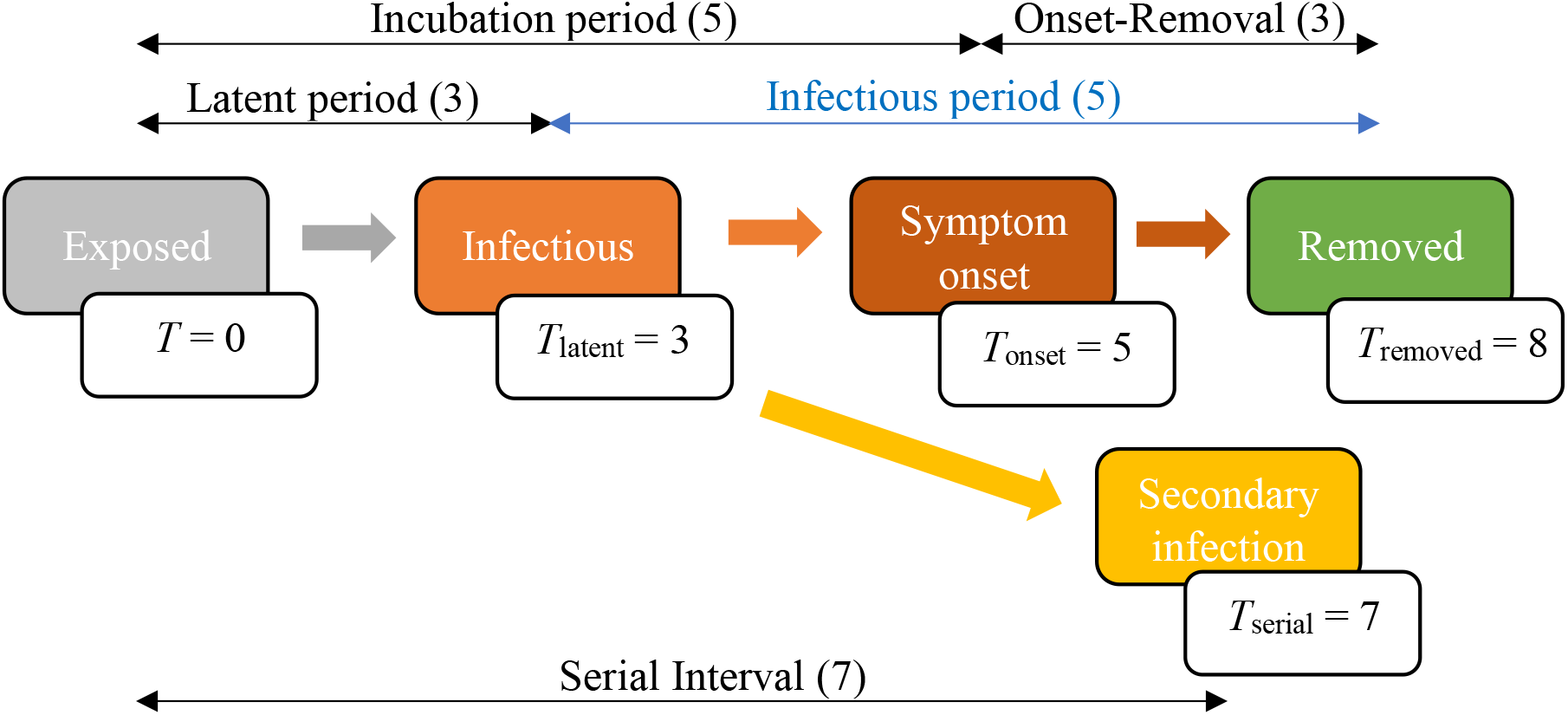
Timeline of infection stages with typical parameter estimates for COVID-19 in Singapore.

The *incubation period* is the time from exposed to the onset of symptoms. The mean incubation period for COVID-19 is estimated to be 5 days [4, 29]. The infectious period is the time between becoming contagious to the time of removal or recovery. Hence, it is the difference between the time of removal and the latent period (*T*_moved_ – *T*_latent_).

In Singapore, the 14-day average time from the onset of symptoms to removal ranges from 1.5 to 6 days after the start of the Circuit breaker on April 7, 2020 (Figure 3).

**Figure 3.**
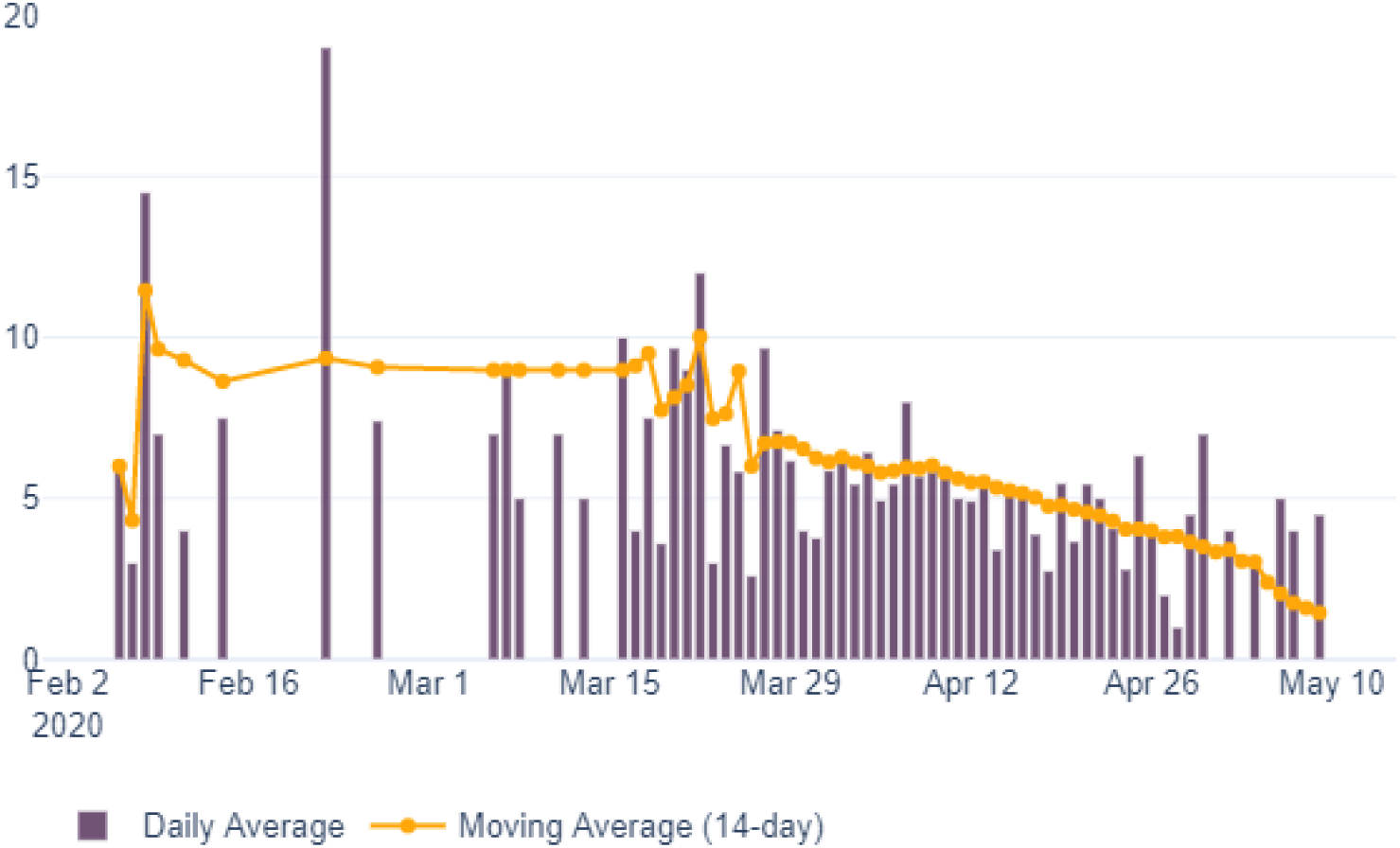
Average number of days from onset of symptoms to isolation for community unlinked cases in Singapore [1].

The serial interval is the time when a secondary infection is generated. For COVID-19 in Singapore, the serial interval between transmission pairs ranges between 3 days and 8 days [5]. Other researchers have reported serial intervals within the same range [6,7,8, 29].

## 3. Method for estimating the basic reproduction number using the SEIR model

One of the important numbers in epidemiology is the basic reproduction number, *R_0_*, defined as the expected number of infections directly generated by one case in a population in which all individuals are susceptible to infection [15, 28]. *R_0_* can be estimated by fitting the data on reported infections to a Susceptible-Exposed-Infectious-Removed (SEIR) model.

Figure 4 is a graphical representation of the classic deterministic SEIR model which shows people passing through four infection states - Susceptible (*S*), Exposed (*E*), Infected (*I*), and Removed (*R*) [12, 13, 14, 27]. The model is governed by four differential equations.

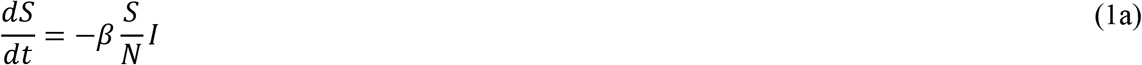

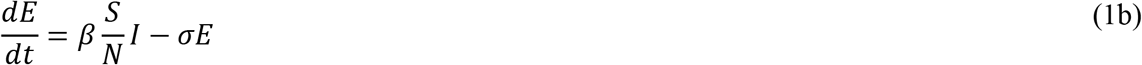

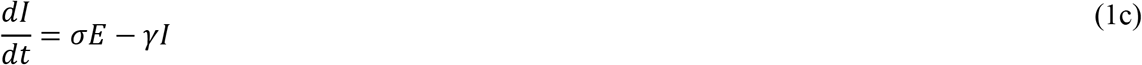

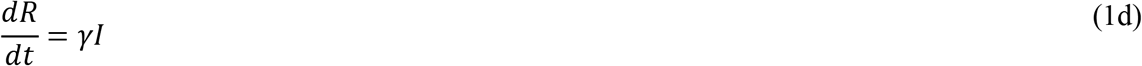

**Figure 4.**
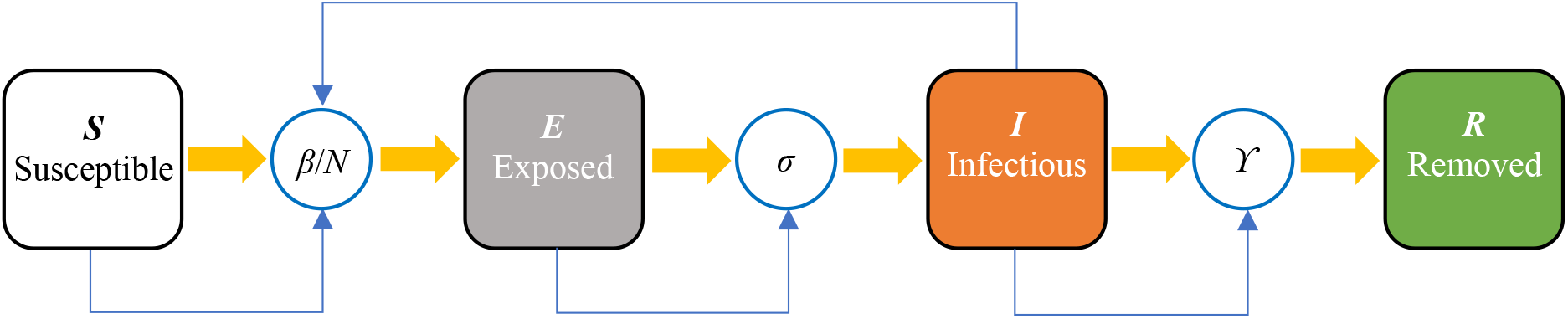
Susceptible-Exposed-Infectious-Removed (SEIR) model of infection.

**Table 1.**
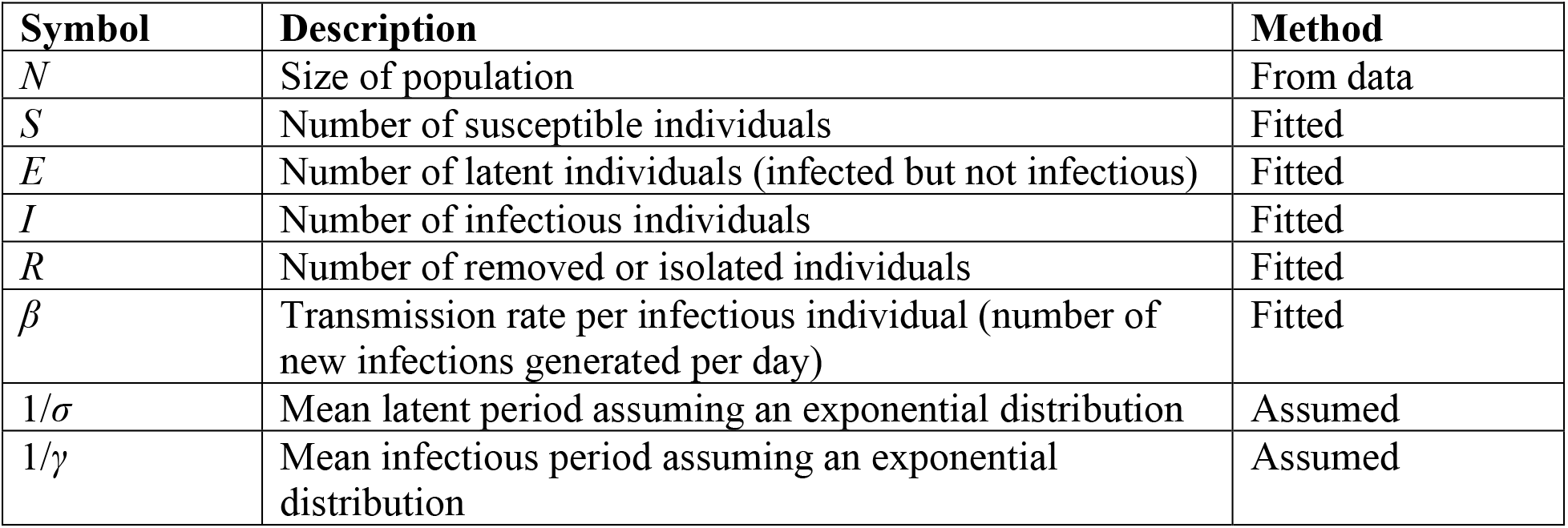
Parameters and initial conditions of the SEIR model

Given a set of initial conditions, the progression from *S*, *E, I*, to *R* over time can be calculated. The differential equations can be solved numerically in Microsoft Excel by using the Runge-Kutta 4^th^ order method. Data on the daily number of reported cases [2, 31] can be compared with the calculated values of the Removed (*R*) class. *R* is defined as the number of removed or isolated individuals and it is calculated according to the SEIR model.

To estimate the model parameters (*S*, *E*, *I*, *R*, and *β*), we use the maximum likelihood method [15]. We assume that the number of reported cases, *x_i_*, at time, *t_i_*, follows the Poisson distribution and has a mean of *μ_i_*, where *μ_i_* is *R*, the calculated number of removed individuals at time, *t_i_*.

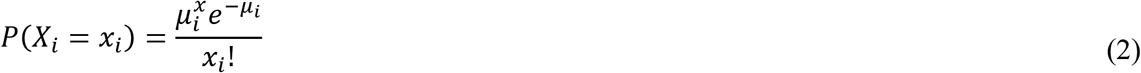

Then, the log-likelihood function to be maximized is

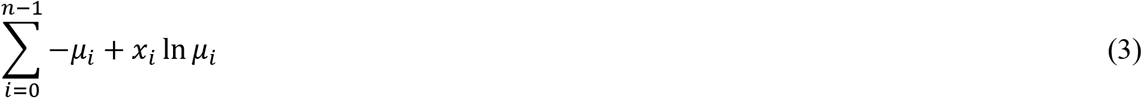

We choose the parameter values for *I*(*t* = 0) and *β* that would maximize the log-likelihood function. This can be done by using the Solver function in Excel. From the SEIR model, the basic reproduction number can then be calculated according to the formula *R*_0_ = *β/γ*.

### Accounting for changing reproduction number

With intervention, specifically social distancing measures, the transmission rate, *β*, and reproduction number will change over time. For a good fit and to obtain a moving average, parameter estimation is best done over a relatively short window of time, say between 7 and 14 days. Figure 5 shows an example where the data is fitted over a 14-day period.

**Figure 5.**
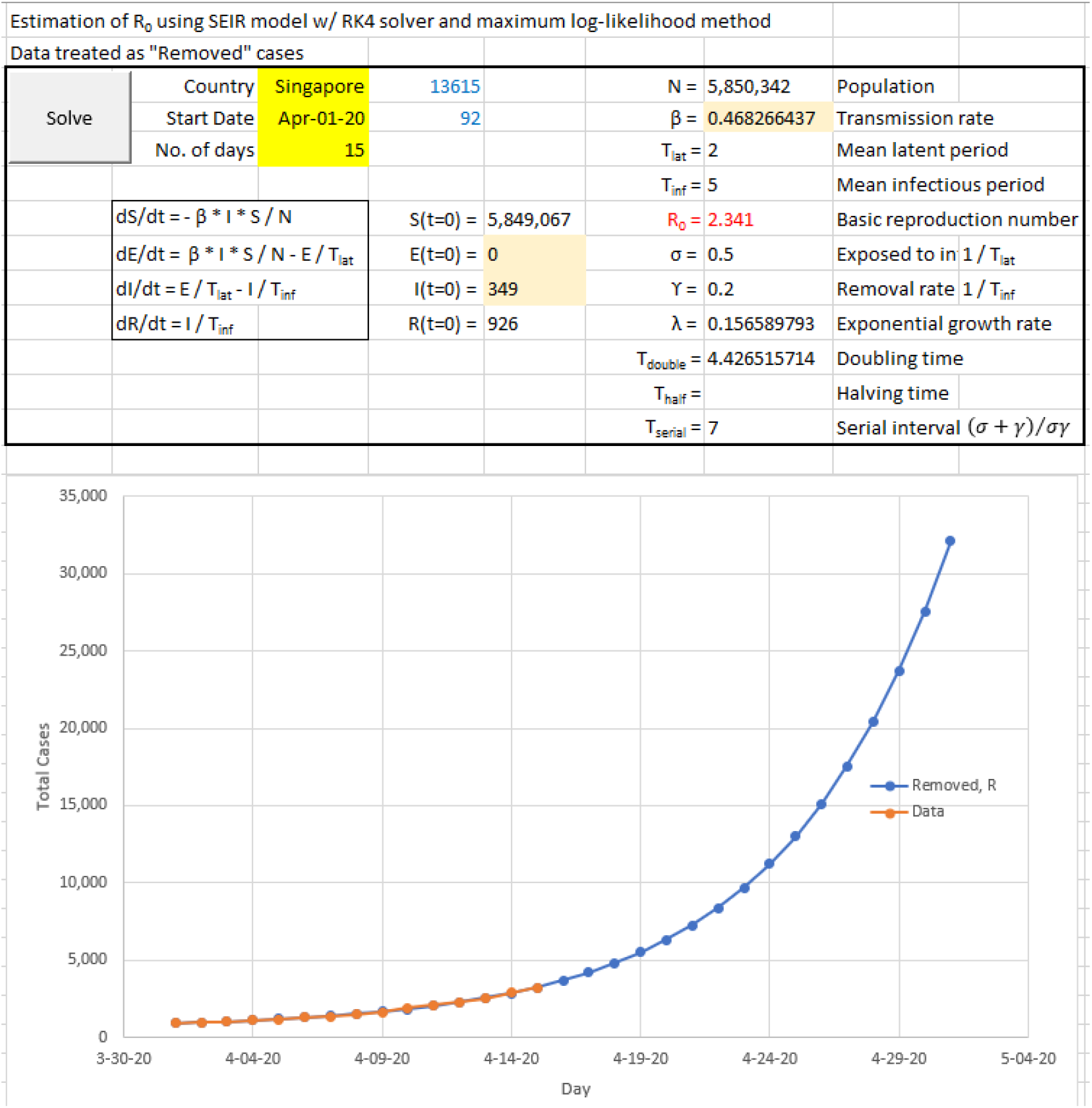
A screenshot of the Excel spreadsheet for estimation of *R_0_*

### Accounting for undetected infection

It is often the case that not all infected cases are reported. To account for undetected infection, one strategy is to estimate the ratio of reported cases (*R*) to total cases (*C*), *p*, and compare this to known values of local case morbidity rate of the disease, *ρ* (adjusted for age distribution and clinical care) [16].

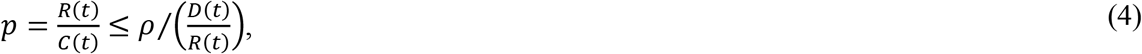

where *D*(*t*) is the reported number of cases that have required critical care or have died. *R*(*t*) is the reported number of cases in any period. Figure 6 provides a visual representation of this concept. If we can assume *ρ* to be constant across regions, then the smallest value of *ρ* across such different regional data may provide an upper bound for *p*. Data from studies have suggested about 5 per cent of COVID-19 patients require critical care [1].

**Figure 6.**
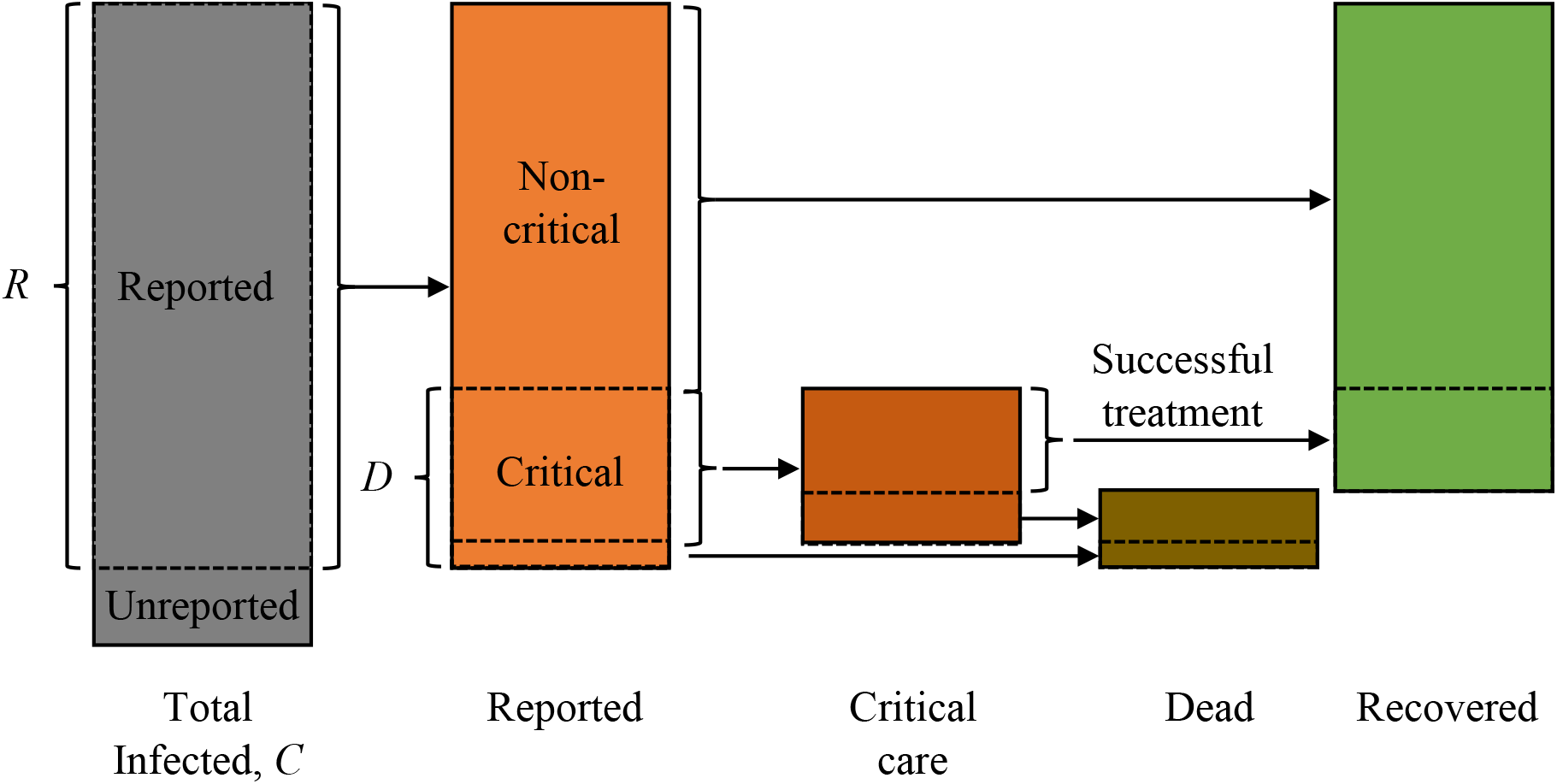
A diagram to illustrate the proportion of different groups at various stages of infection

In a test sample of the Singapore population, 39,000 preschool staff were screened and 13 tested positive using the polymerase chain reaction (PCR) test [1]. This represents about 0.035 per cent of those tested. Hence, the number of undetected infections in Singapore is currently considered small.

## 4. Method for deriving symptom onset dates from confirmation dates

The daily number of reported cases is partly dependent on the number of tests conducted, which may be variable due to factors such as testing capacity and the day of week. To account for this variation, we perform a running 3-day average of test cases. Other methods of applying a smoothing filter to the time series may be used if appropriate.

Another issue is the delay between the onset of symptoms and case confirmation (removal or isolation). Case onset dates can be derived if records of onset-to-confirmation dates are available for every individual (e.g. see Fig. 2). Otherwise, case onset dates can be estimated by using the following procedure.

i. For each date, distribute case counts back in time according to a Poisson distribution with a mean of 3 days (symptom onset to removal) as illustrated in Figure 7.
ii. Sum the back distributed case counts for each date to derive the onset curve as shown in Figure 8.
iii. Distributing reported cases back in time and recreating the onset curve result in a “right-censored” time series. This means that there are onset cases close to the present date that are yet to be reported. We correct this by estimating the percentage of onset cases on Day (*t-a*) that have not yet been reported by today (Day *t*). We can use the cumulative distribution function of the Poisson “onset-to-removed” distribution to adjust for the number of onset cases, thus removing right censoring.

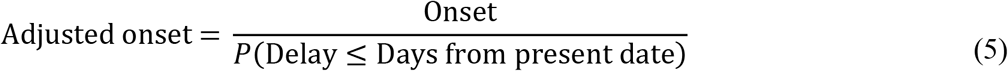 Consider an example illustrated in Figure 9. Three days ago, there were 470 reported onset cases. This represents the fraction of the actual number reported over the next 3 days. This fraction is equal to the value of the cumulative distribution function of our Poisson distribution at Day 3, which is 65%. Hence, the current count of onset on that day represents 65% of the actual total. After adjustment, the actual total is estimated to be (1/0.65) of 470, which is 723. Figure 10 shows the adjusted onset curve.

**Figure 7.**
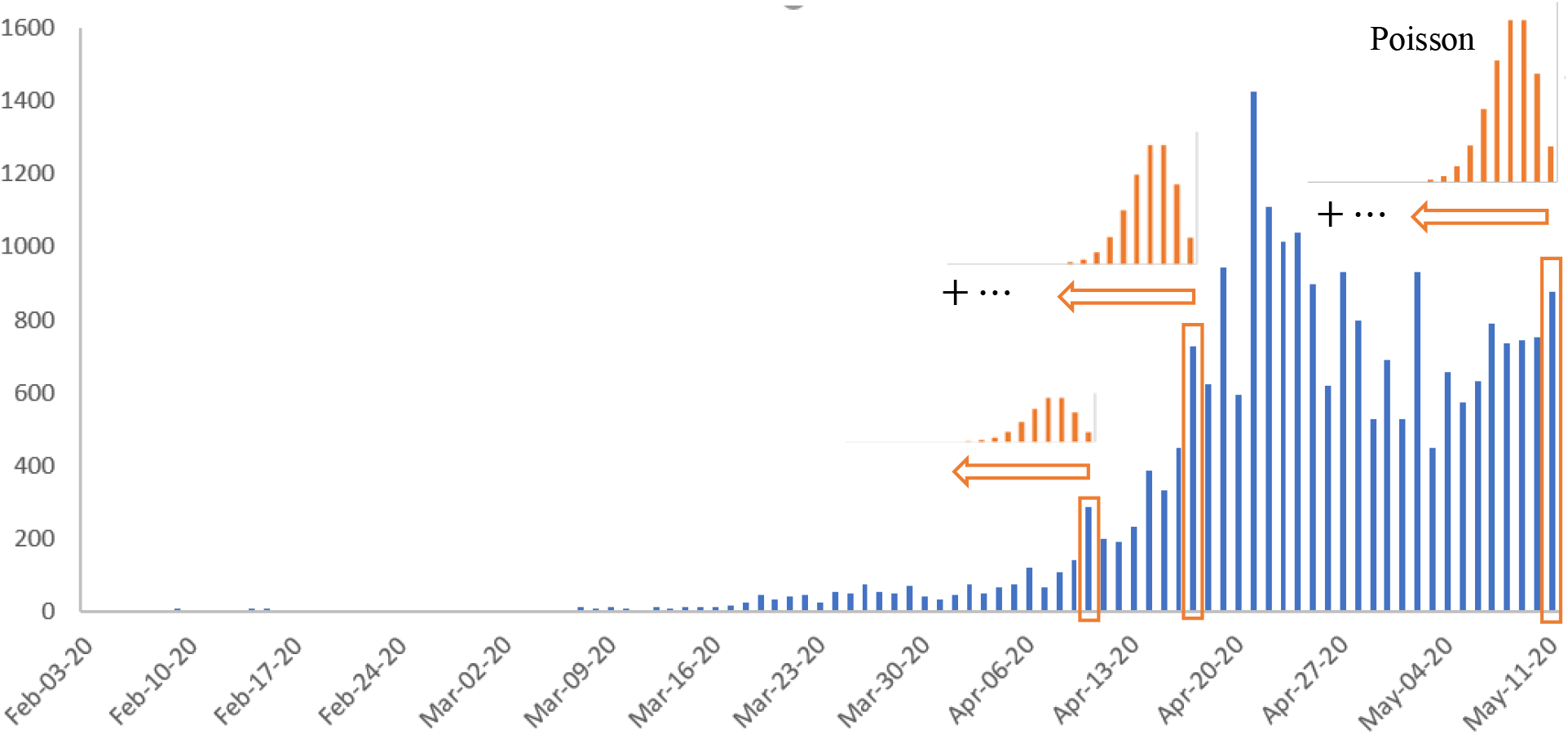
Distributing case counts back in time.

**Figure 8.**
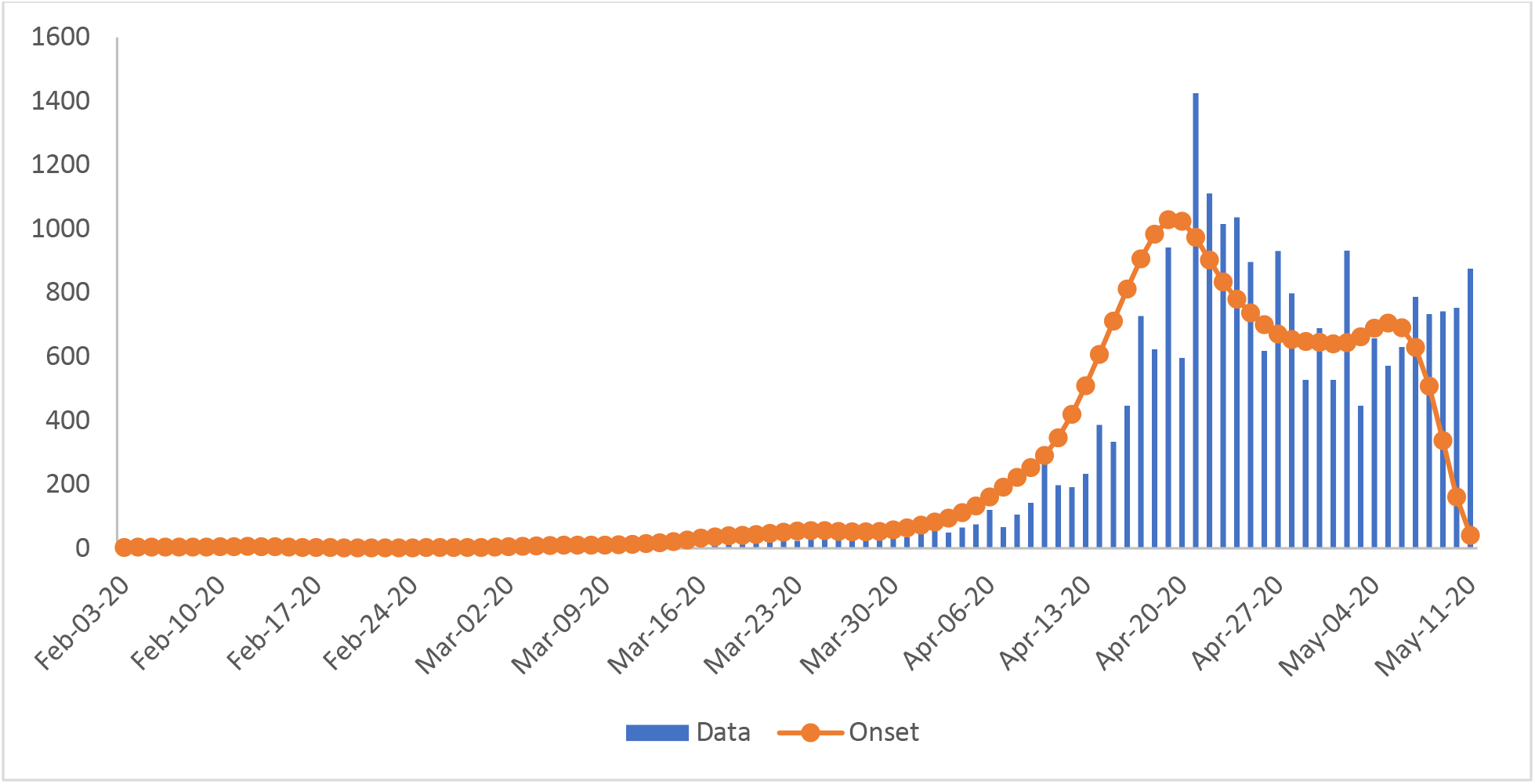
The onset curve estimates the cases during the onset of symptoms.

**Figure 9.**
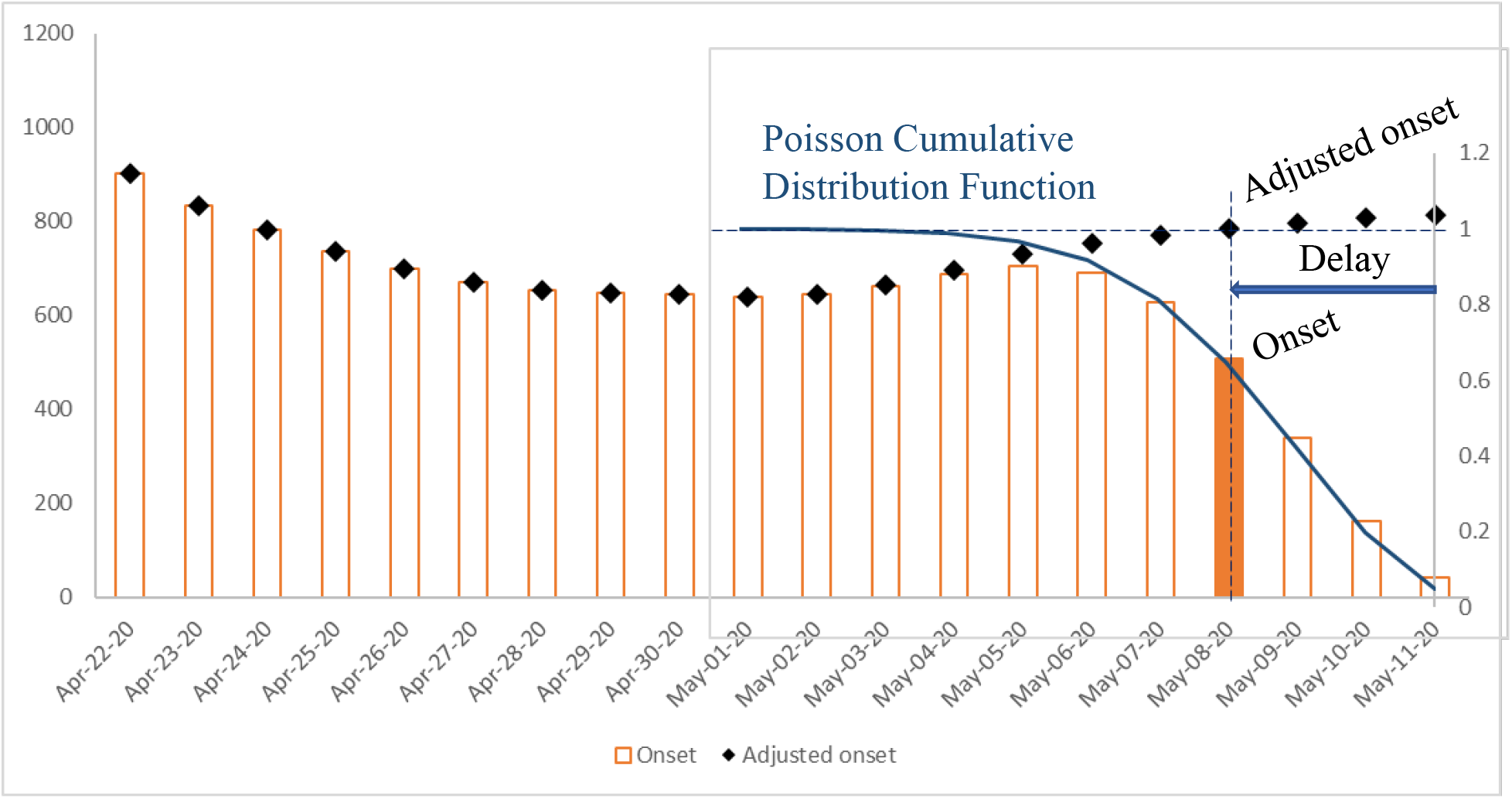
Adjusting for right-censoring.

**Figure 10.**
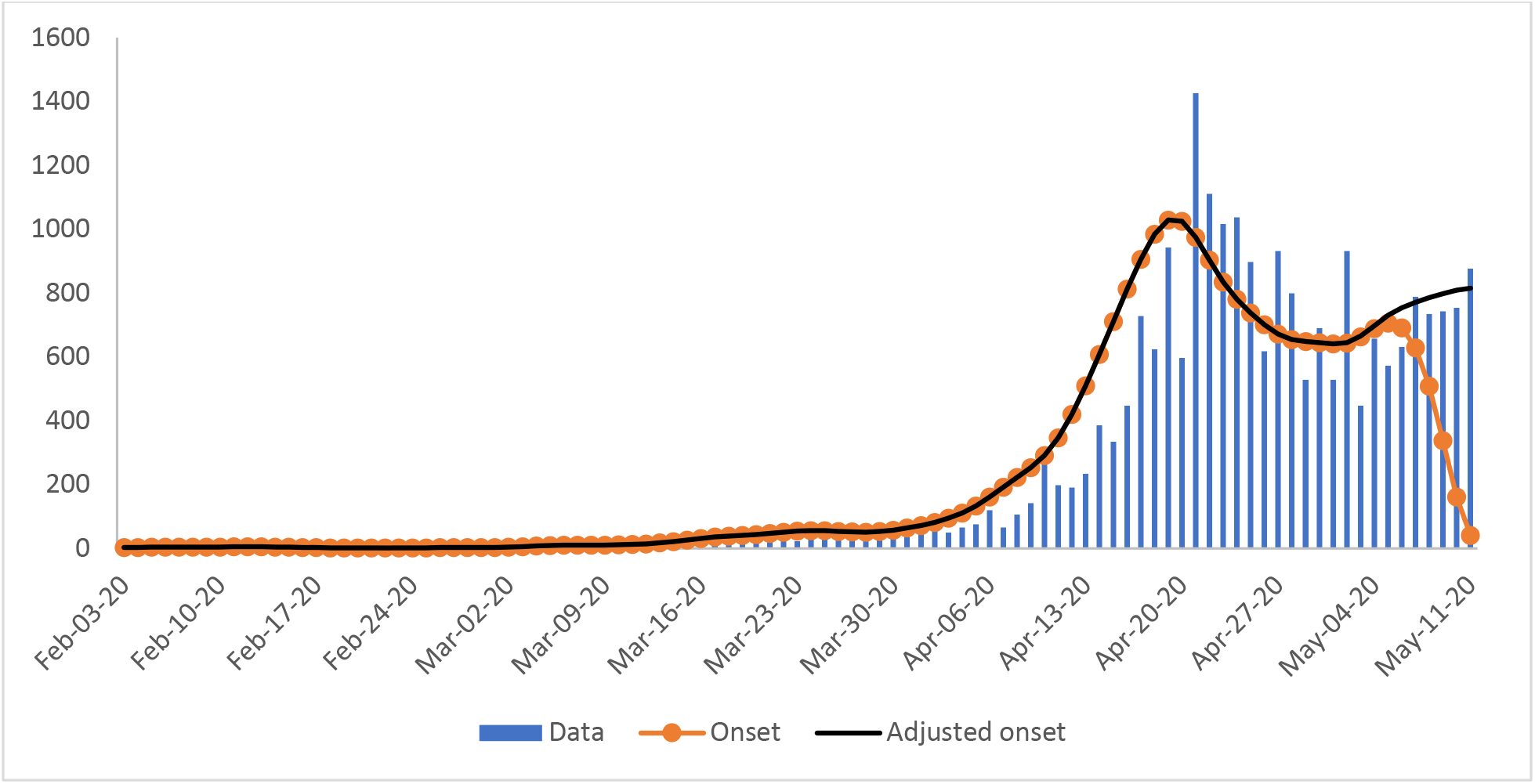
Onset numbers close to the present date are adjusted for right censoring.

## 5. Method for deriving infection (exposed) dates from onset dates

A similar procedure as in Section 4 can be applied to the onset counts to derive the infection (exposed) time series. Figure 11 shows the adjusted exposed time series where the incubation period (from exposed to symptom onset) follows a Poisson distribution with a mean of 5 days.

**Figure 11.**
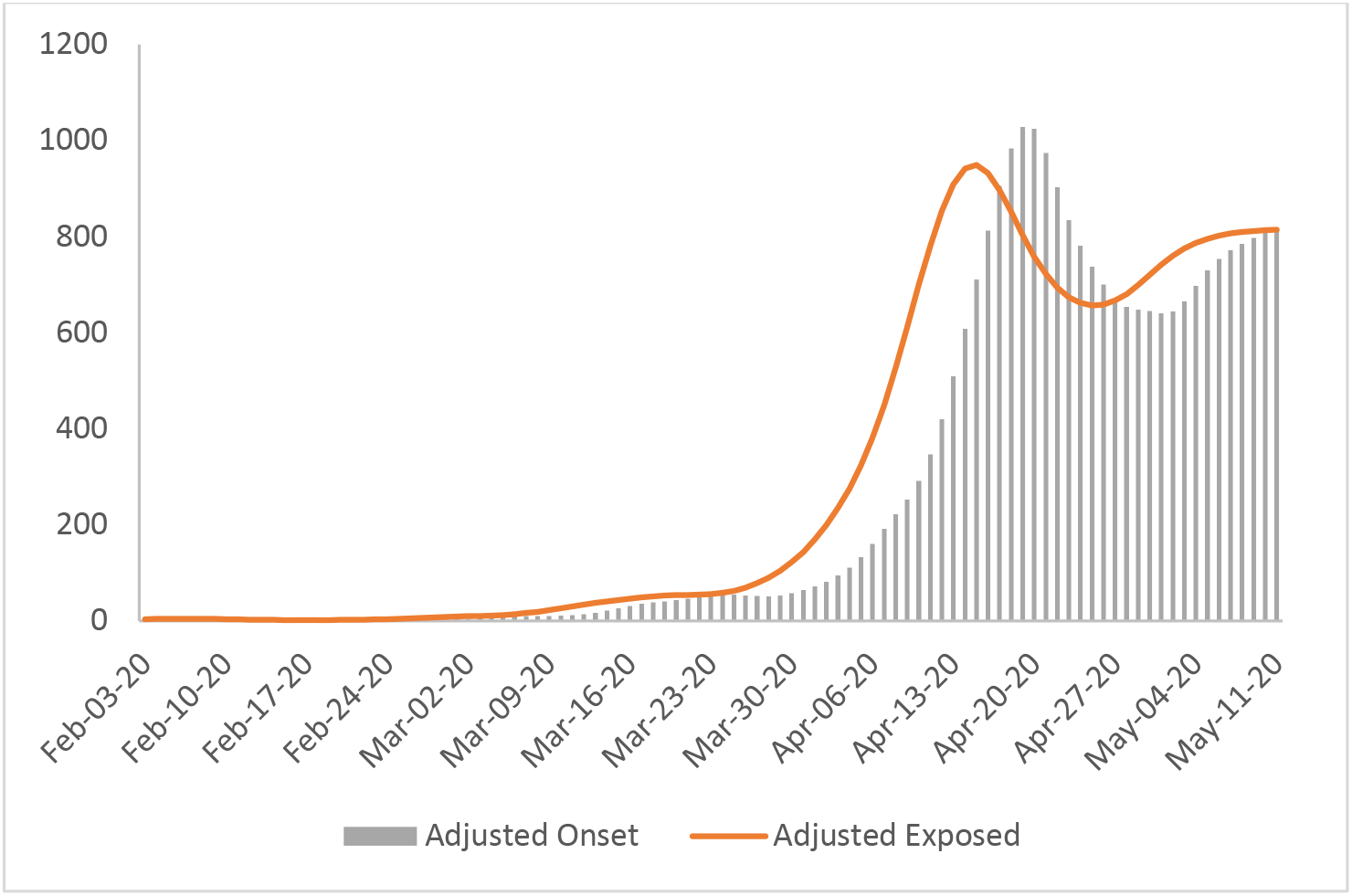

## 6. Method for estimating the effective reproduction number, *R*(*t*)

The basic reproduction number, *R*_0_, is the expected number of infections directly generated by one case given that all individuals are equally susceptible. As the infection spreads, the susceptibility of the population decreases. The effective reproduction number, *R*(*t*), is related to the basic reproduction number, *R*_0_, by *R*(*t*) = *R*_0_*S*(*t*), where *S*(*t*) is the average susceptibility of the population. *R*(*t*) is often used as an indicator of the effectiveness of interventions, such as social distancing measures, to contain the spread of a virus. If *R*(*t*) is greater than 1.0, the infection is growing at an exponential rate. If *R*(*t*) is at 1.0, the spread is sustained at a linear rate. If *R*(*t*) is less than 1.0, the infection is spreading at a slower pace and will eventually die out.

Although *R*(*t*) cannot be measured directly, it can be estimated via two methods. We describe these two methods that can be implemented in a spreadsheet without any programming codes.

### 6.1 Non-parametric approach

Wallinga and Lipsitch [17, 18] have developed a non-parametric method to derive the reproduction number from the exponential growth rate,

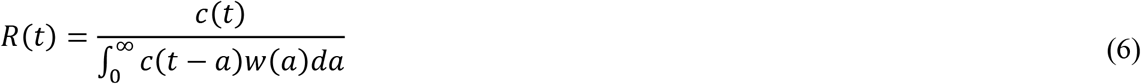

where *c*(*t*) is the rate of new infections at time *t*, and *w*(*a*) is the probability density function of the serial interval (see Figure 2).

The serial interval is the time from being infected to generating a secondary infection. We assume a gamma distribution with a mean serial interval of 7 days and a peak (most infectious) at Day 4 (Figure 12). This accounts for a latent period where the exposed individual is not yet infectious.

**Figure 12.**
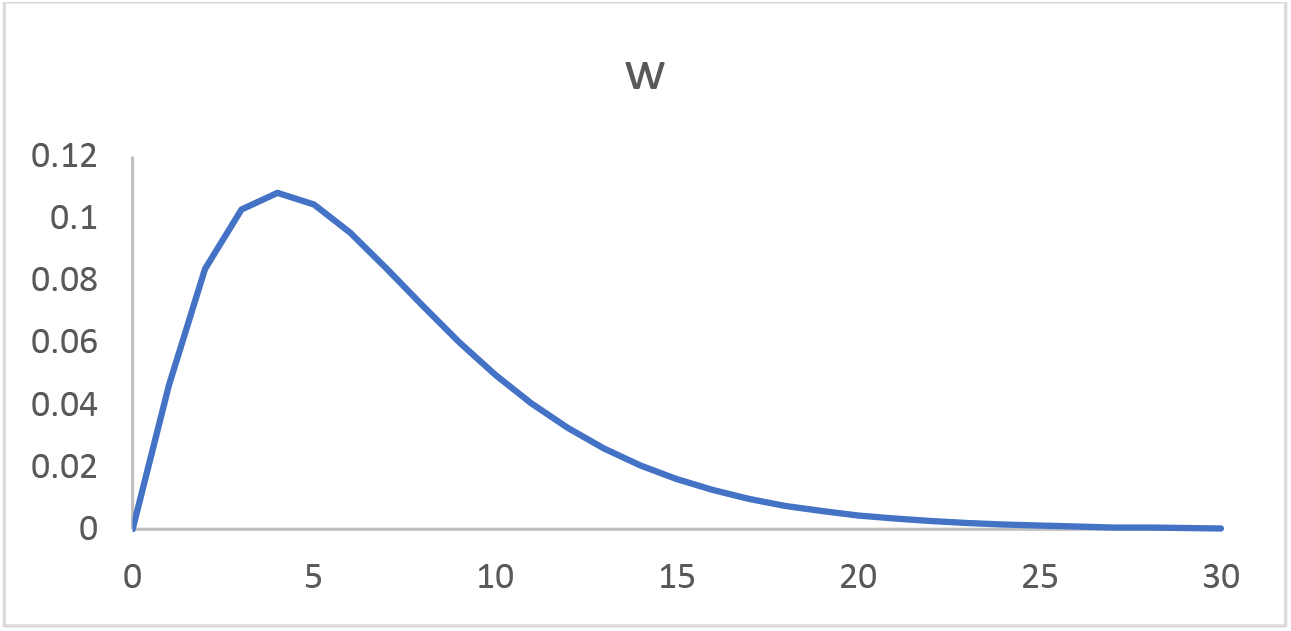
A gamma distribution calculated using the GAMMA.DIST function in Excel.

Equation (6) can be evaluated in a spreadsheet using the infection data derived above and a numerical integration scheme. Figure 13 shows a plot of *R_t_* against time for Singapore during the Circuit breaker period.

**Figure 13.**
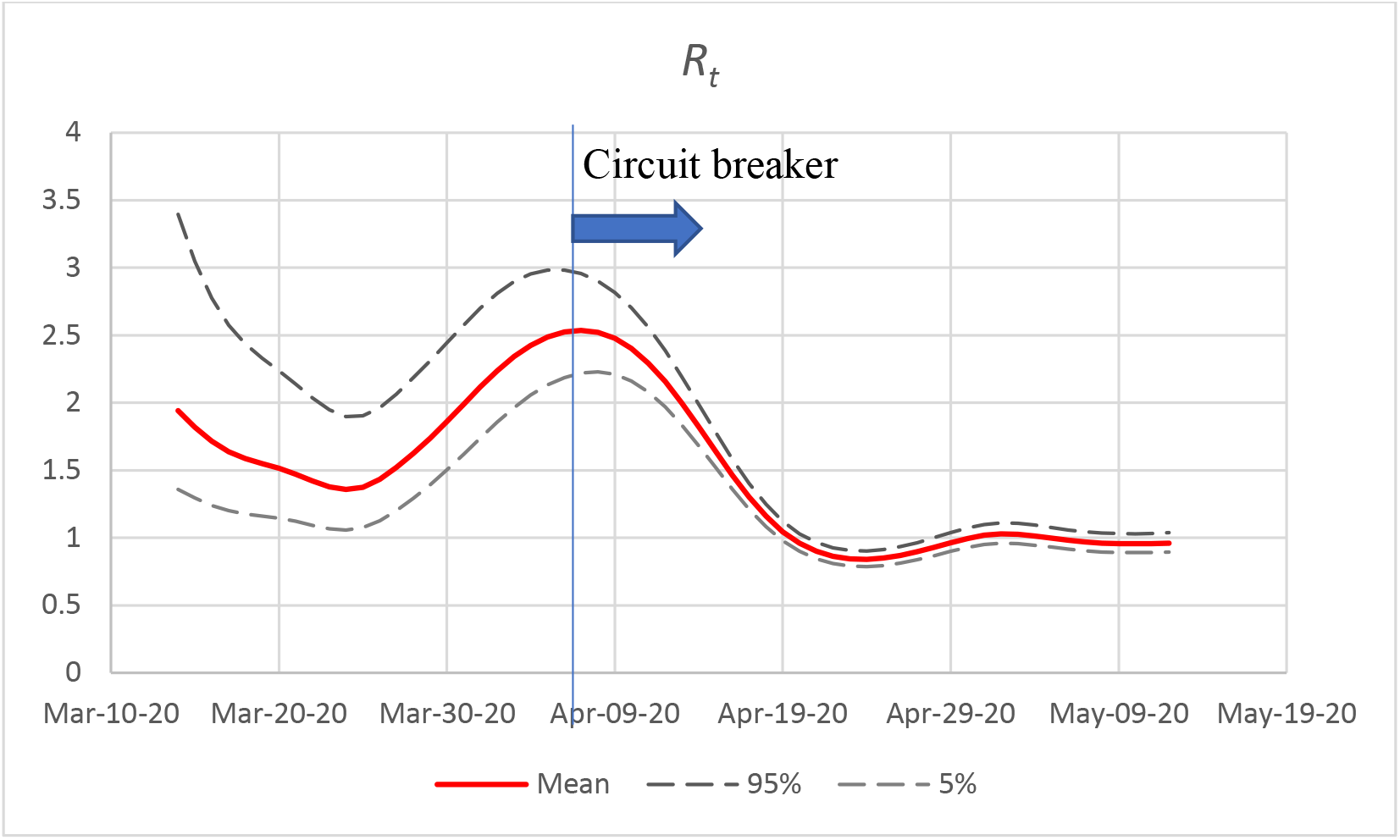
*R_t_* against time for Singapore during the Circuit breaker period

The results clearly show that the Circuit breaker measures imposed from April 7, 2020 have an immediate effect of rapidly slowing down the spread of COVID-19. We can also see that *R_t_* settled to around 1.0 after about two weeks. Since then, the infection rate has remained sustained. Given that dormitory residents make up the majority of the infected individuals, it can be concluded that individuals continue to infect others with a reproductive ratio of approximately 1 to 1 in that setting during that time period as depicted in Figure 13.

To estimate the confidence intervals of the *R*(*t*) estimate, we first discretize equation (6),

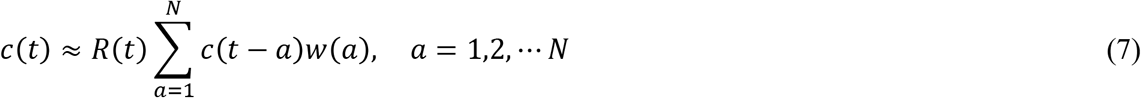

Note that the expected number of secondary infections generated on Day *t* by individuals infected on Day (*t-a*) is

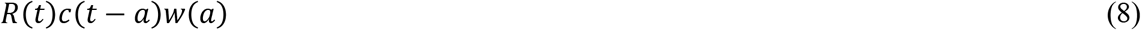

where *c*(*t-a*) is the number of new infections *a* days before the present day, and *w*(*a*) is the probability that the serial interval is *a*. Not all the *c*(*t-a*) cases will generate new infections on Day t. In general, the probability that *k* out of *n* cases will generate secondary infections with a serial interval of *a* is binomially distributed,

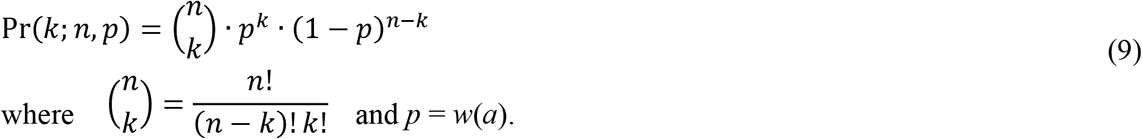

This is a binomial distribution with a mean of *nw*(*a*) and a variance of *nw*(*a*)(l − *w*(*a*)). Table 2 shows the formulas for the probability distributions for *N* days before Day *t* (the present date).

**Table 2.** Formulas for probability distributions for *N* days

| $a$ | Day | Probability that serial interval is $a$ | No. of infected individuals on Day $(t-a)$ | Probability of having $k$ individuals with serial interval $a$ |
| --- | --- | --- | --- | --- |
| 1 | $t-1$ | $w(1)$ | $c(t-1)$ | $\binom{c(t-1)}{k_1} \cdot w(1)^{k_1} \cdot (1-w(1))^{c(t-1)-k_1}$ |
| 2 | $t-2$ | $w(2)$ | $c(t-2)$ | $\binom{c(t-2)}{k_2} \cdot w(2)^{k_2} \cdot (1-w(2))^{c(t-2)-k_2}$ |
| ⋮ | ⋮ | ⋮ | ⋮ | ⋮ |
| $N$ | $t-N$ | $w(N)$ | $c(t-N)$ | $\binom{c(t-N)}{k_N} \cdot w(N)^{k_N} \cdot (1-w(N))^{c(t-N)-k_N}$ |

The probability for the total number of secondary infections on Day *t*, 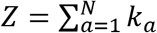, is the sum of *N* binomial distributions. Thus, using the binomial sum variance inequality [19, 20], the upper bound of the variance of *Z* can be calculated.

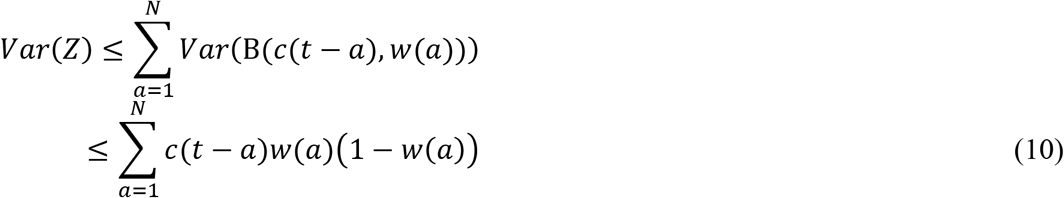

Equation (10) allows us to estimate the confidence intervals of the *R*(*t*) estimates.

### 6.2 Bayesian approach

The Bayesian approach allows us to continuously update our estimate of a set of parameters, Θ, as more data becomes available.

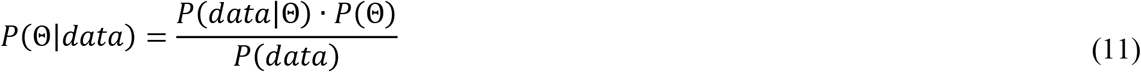

*P*(Θ), the prior distribution, represents our prior estimates about the true value of Θ.

*P*(*data*|Θ) is the likelihood distribution. It is also often written as ℒ (Θ |*data*) which means the probability of observing the data given Θ. For the method to work, it is necessary to calculate the likelihood distribution for all possible values of Θ.

*P*(*data*) is the model evidence and it is the same for all possible hypotheses (values of Θ) being considered.

*P*(Θ|*data*) is the posterior distribution and represents our updated estimate of the value of Θ given the observed *data*.

The main objective of Bayesian inference is to calculate the posterior distribution of our parameters using our prior beliefs updated with our likelihood. From the posterior distribution, we can determine the most likely values of Θ given the observed data. Since we are usually only interested in relative probabilities of different hypotheses, *P*(*data*) can be left out of the calculation and we write the model form of Bayes’ theorem as

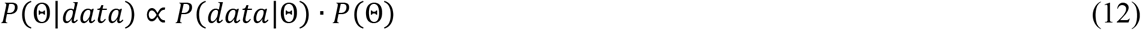

where ∝ means “proportional to”. For estimating *R_t_*, the Bayes’ theorem that we use is

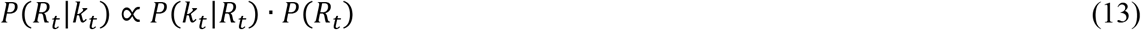

where the data, *k_t_*, is the daily number of cases, and the parameter, *R_t_*, is the effective reproduction number.

Equation (13) is updated every day by using yesterday’s posterior, *P*(*R_t_*_−1_|*k_t_*_−1_), to be today’s prior *P*(*R_t_*). On day two, the equation becomes

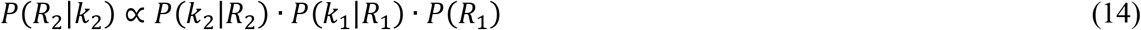

So generally,

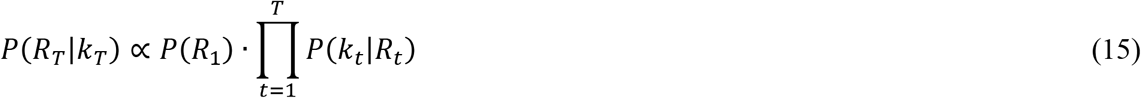

Assuming a uniform starting prior *P*(*R*_1_), this reduces to:

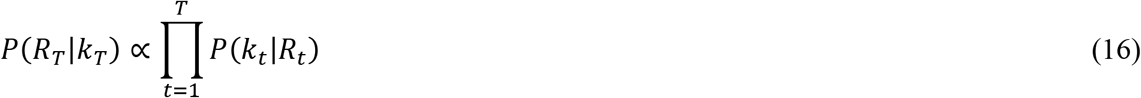

Note that the posterior on any given day is equally influenced by the distant past as much as the recent day. This is fine if we are estimating a static parameter that does not change with time. However, the value of *R_t_* is dynamic and is more closely related to recent values than older ones. To address this issue, we can adopt Systrom’s approach [21] of only incorporating the last *m* days of the likelihood function:

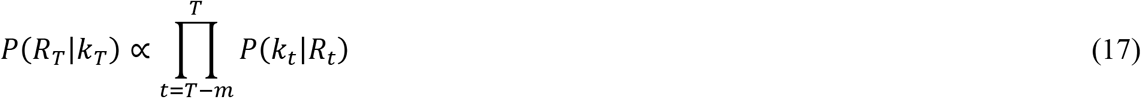

Bettencourt & Ribeiro’s likelihood function

To calculate the likelihood function ℒ(*R_t_|k_t_) = P*(*k_t_*|*R_t_*), we first assume that the number of new infections on any given day can be described by a Poisson probability distribution with a mean of *λ*. The probability of seeing *k* new cases is

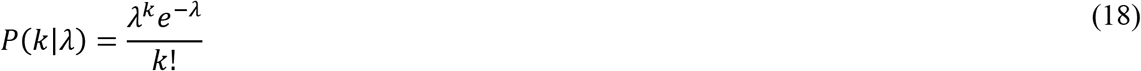

Bettencourt & Ribeiro [22] has derived an equation relating *R_t_* to *λ*.

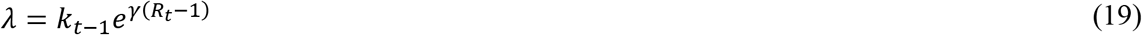

where *γ* is the reciprocal of the serial interval (see Figure 1). Figure 14 shows the variation of *λ* with *R_t_* for some values of *k_t_*_-1_.

**Figure 14.**
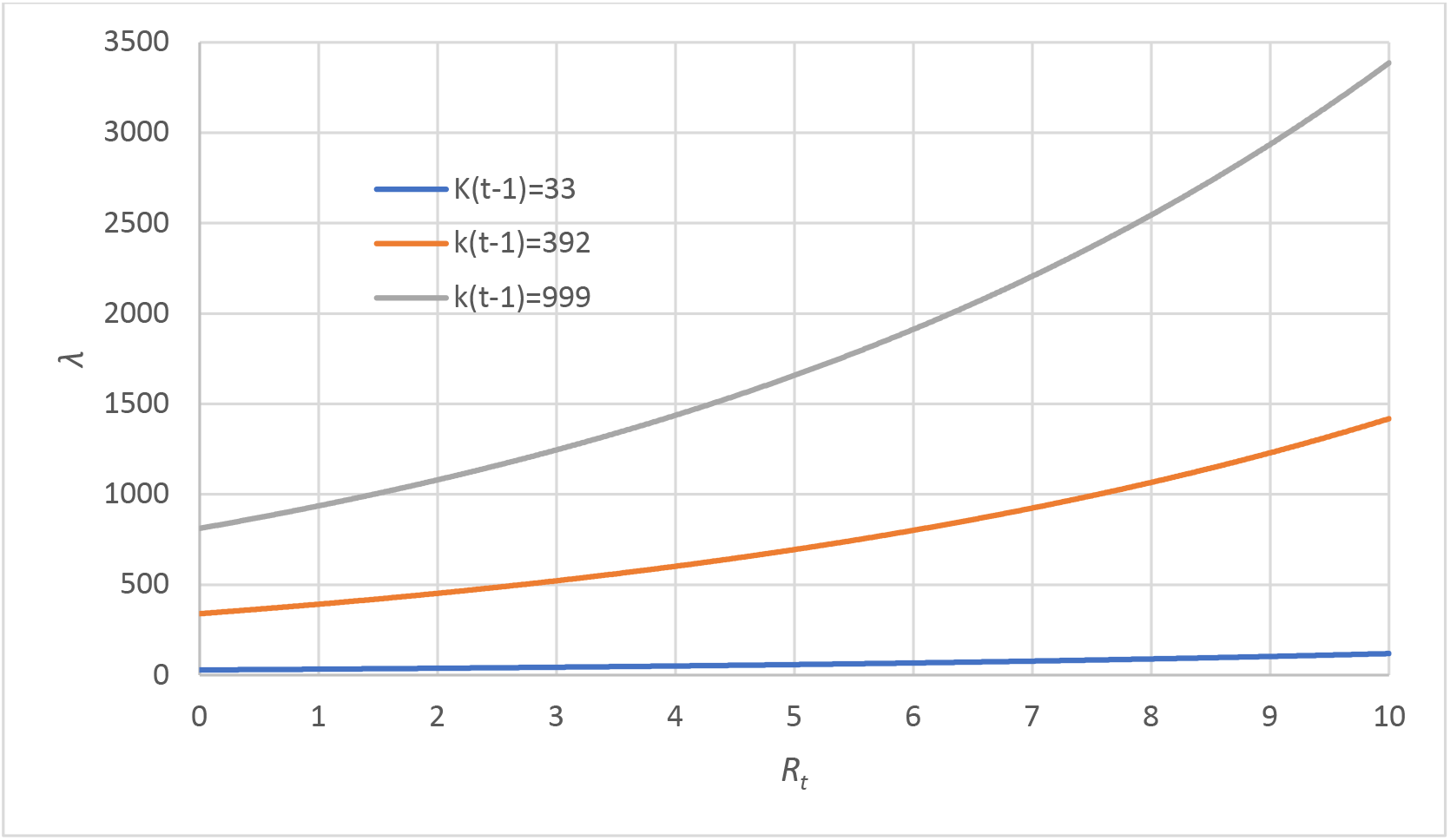
Variation of *λ* with *R_t_* given *k_t_*_-1_.

Equations (18) and (19) allow us to reformulate the likelihood function as a Poisson distribution, parameterized by fixing *k* and varying *R_t_*.

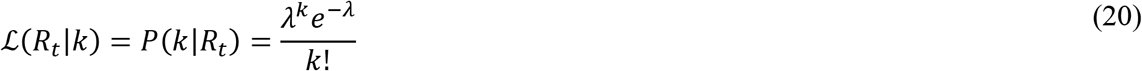

Figure 15 shows that as k increases, the peak value of the likelihood function ℒ(*R_t_|k*) increases and the distribution becomes less spread out about. This means that as the number of infections increases the confidence of our *R_t_* estimate should improve.

**Figure 15.**
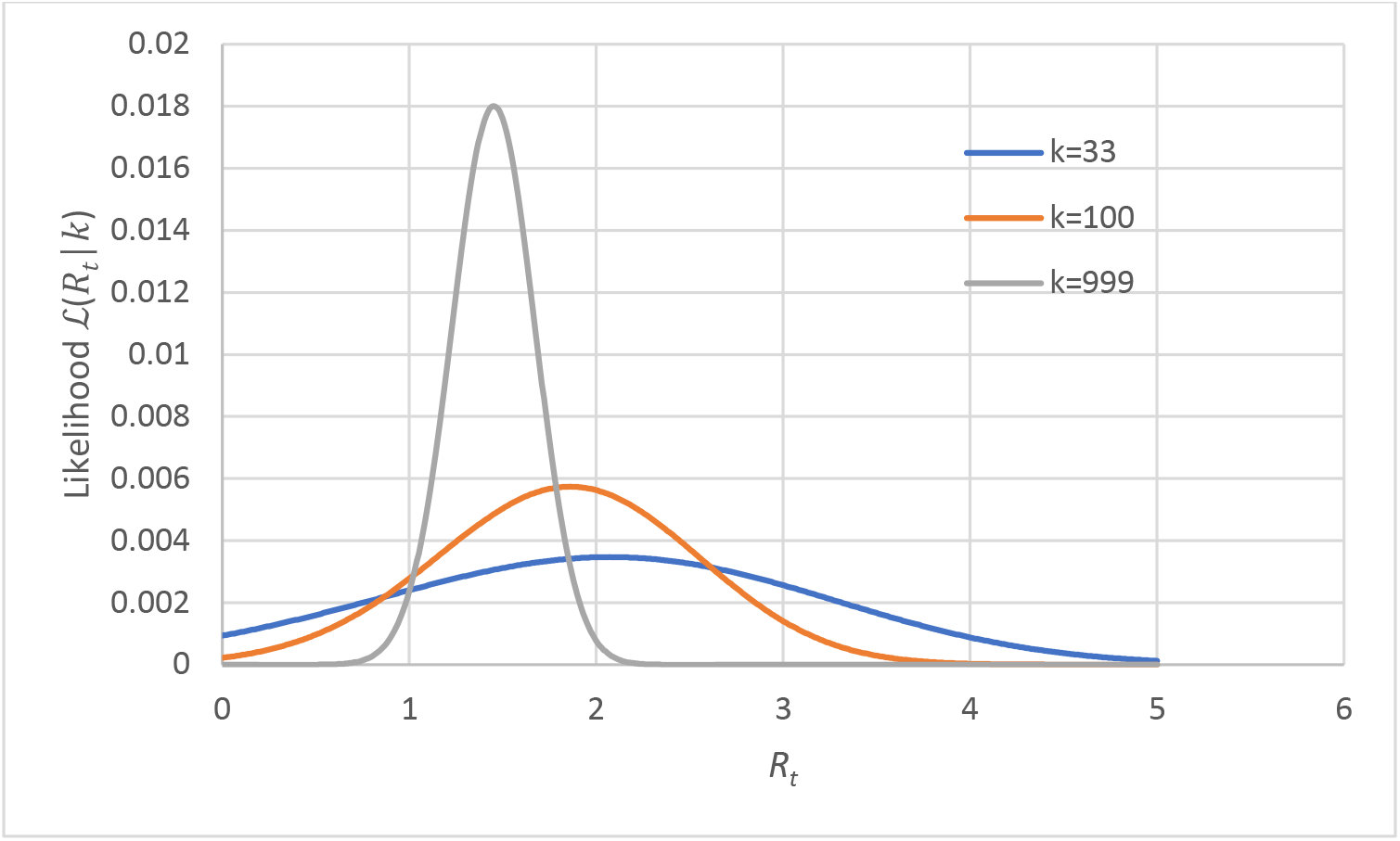
Variation of ℒ(*R_t_|k*) with *R_t_* given *k*.

In evaluating the posteriors, it is more convenient to use the logarithm of the likelihood function.

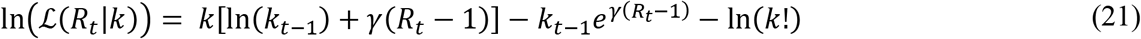

To perform the Bayesian update, we can do a sum of the log-likelihoods over the last *m* days and then exponentiate to get the likelihood. From equations (17) and (21),

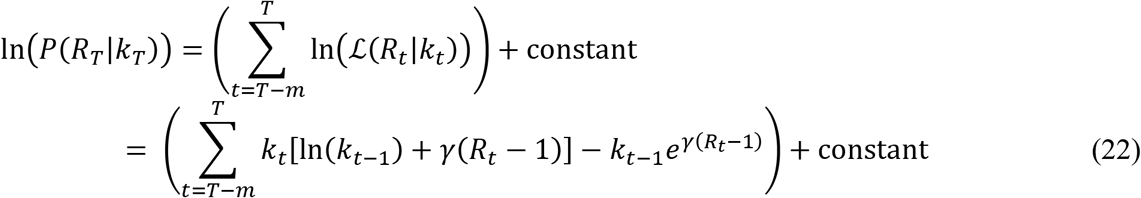

From the posterior distribution (Figure 16) we can also obtain the confidence interval for *R_t_*. For example, Figure 17 shows the most likely values of *R_t_* and the confidence interval over time for Singapore. We can see that *R_t_* changes with time and the confidence interval narrows with more data.

**Figure 16.**
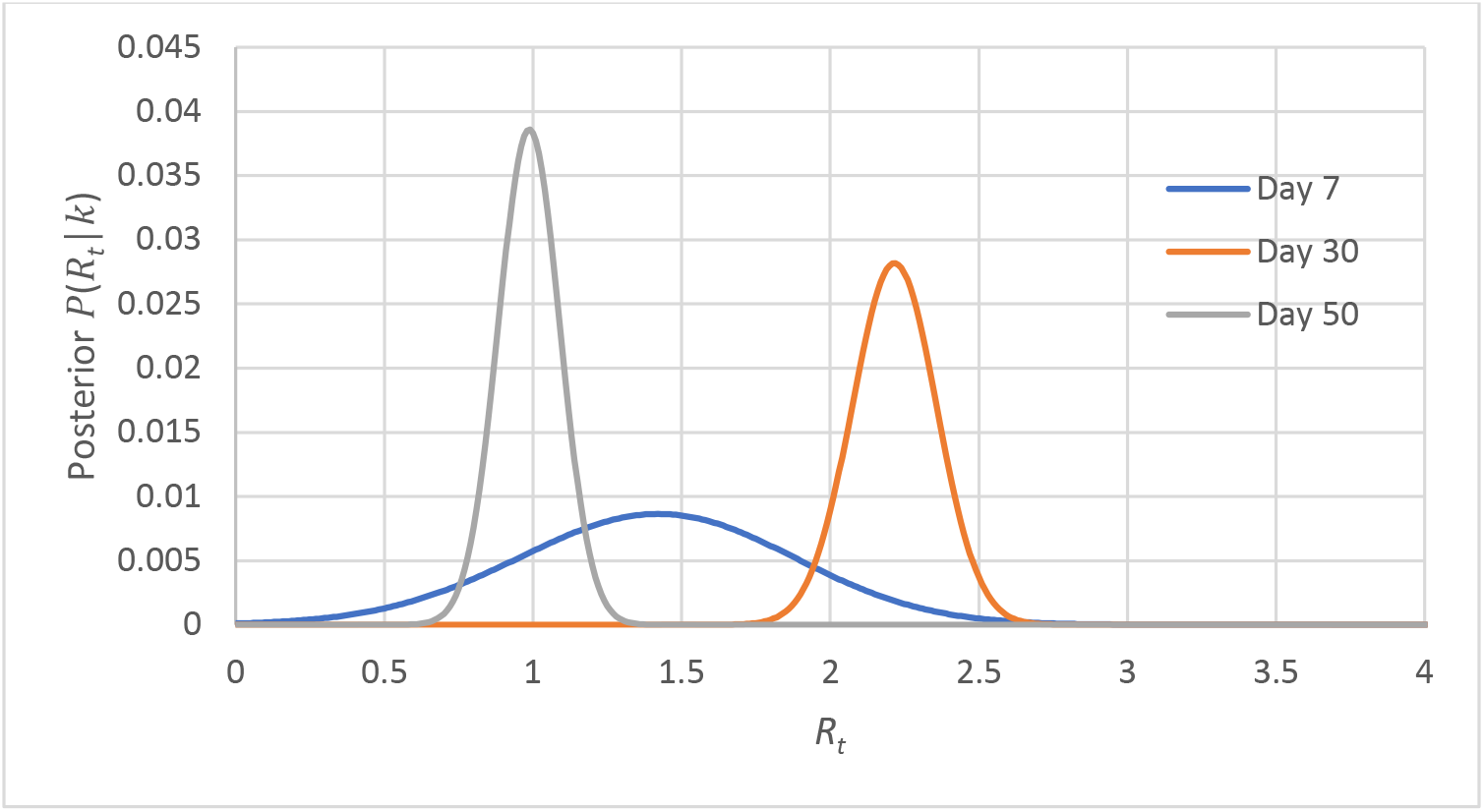
Variation of posterior *P*(*R_t_*|*k*) with *R_t_*.

**Figure 17.**
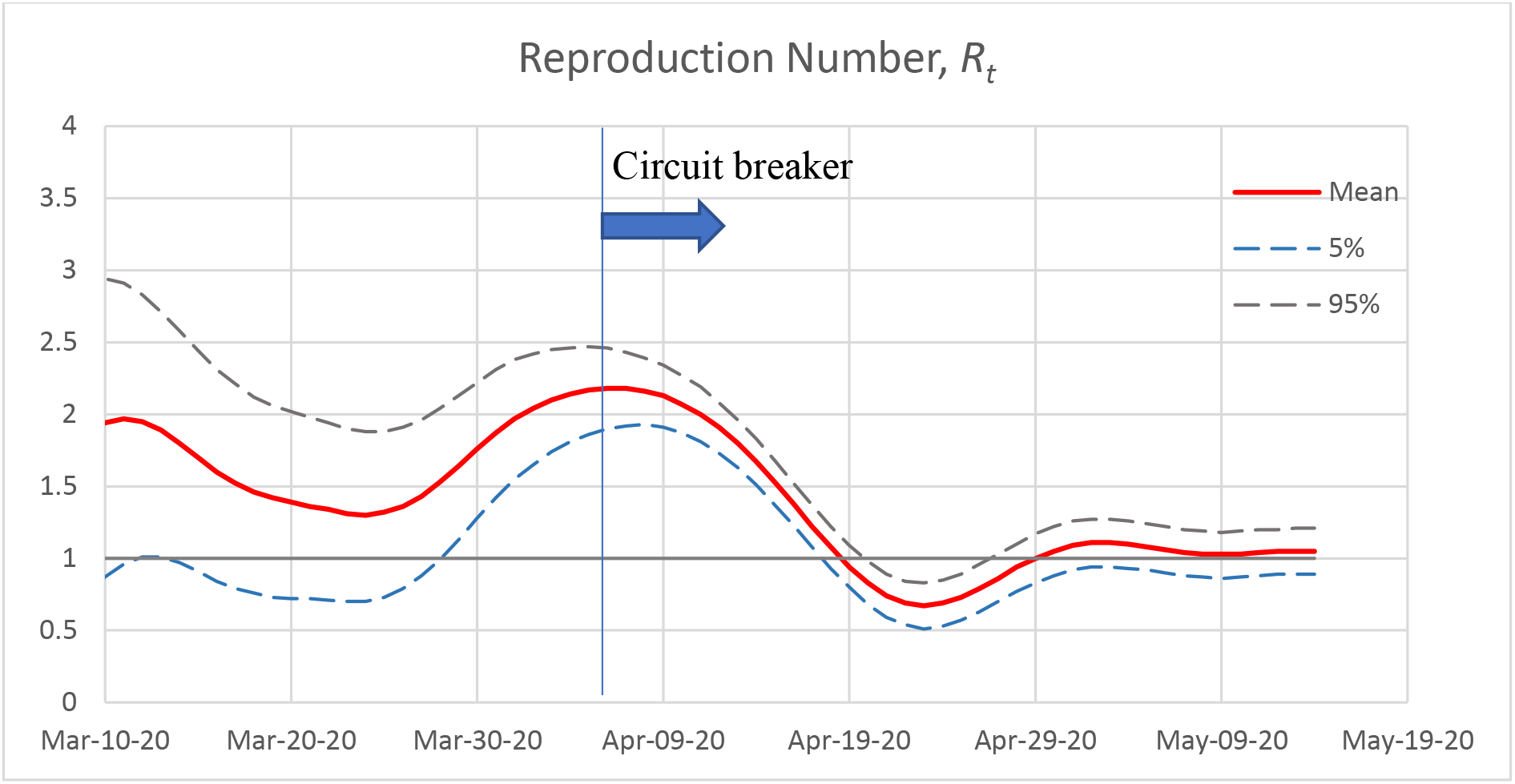
Effective reproduction number *R_t_*.

The results are in agreement with those calculated using the non-parametric approach (Figure 13) and the EpiEstim code (Figure 18) [23, 24].

**Figure 18.**
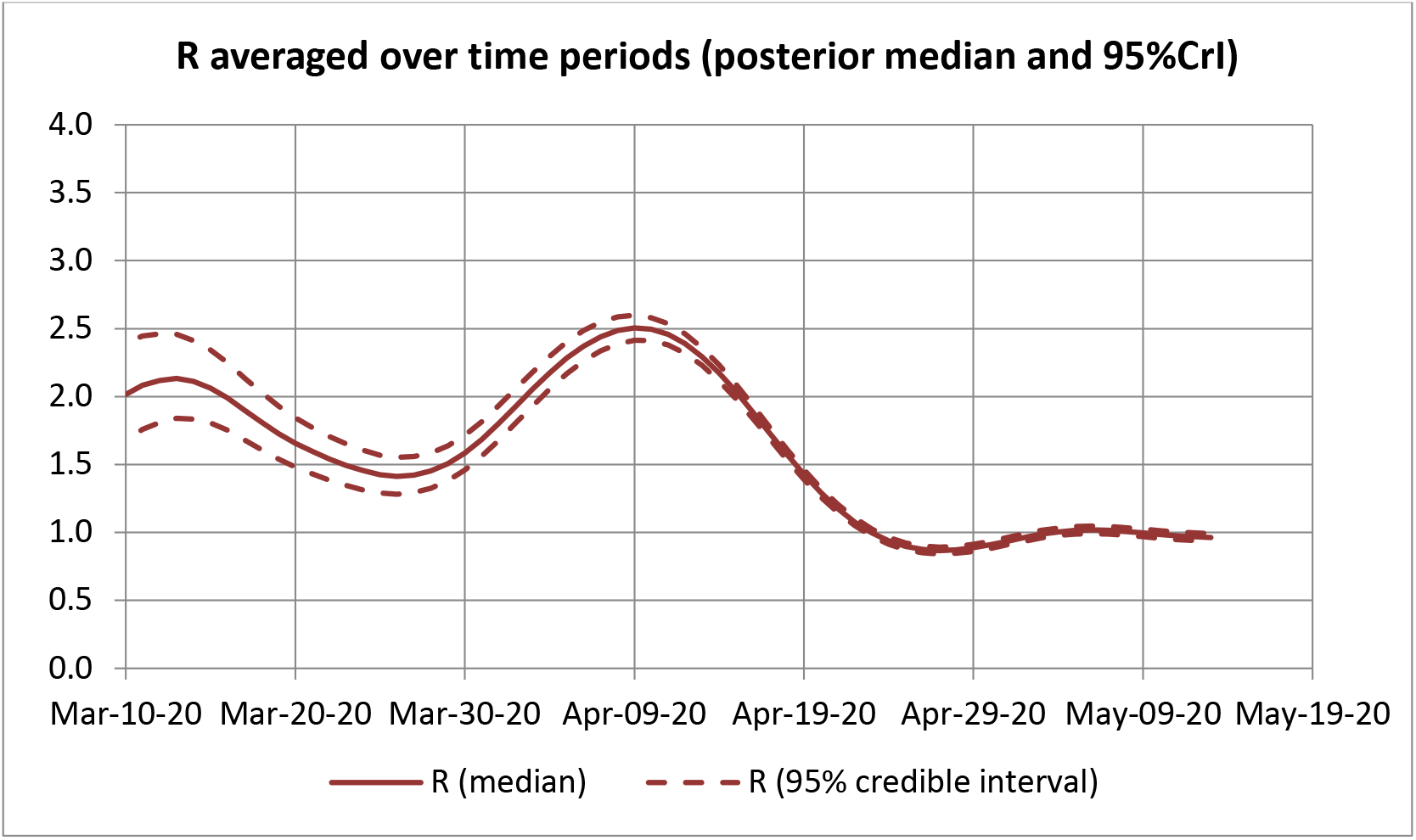
Results from EpiEstim.

## 7. Method for estimating doubling time

Another metric often used to measure infection rate in a population is the doubling time, defined as the time for cumulative cases to double based on the current growth rate [25].

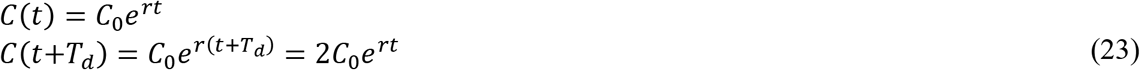

where *C*_0_ is the initial number of cases, *T_d_* is the doubling time, and *r* is the exponential growth rate. Taking the logarithm on both sides of the equation,

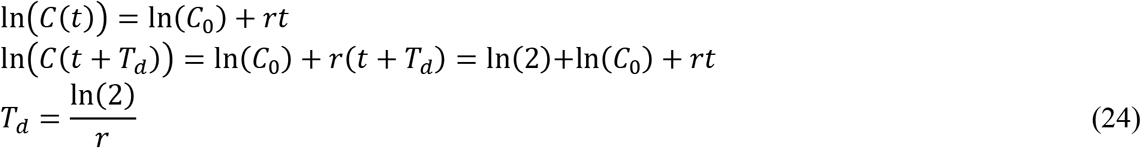

As the growth rate slows, the doubling time increases accordingly. Note that the gradient of the ln(*C*(*t*)) curve is equal to *r*. Time-varying estimates of the doubling time can be made with a 7-day sliding window by iteratively fitting a linear regression model to ln(*C*(*t*)). Figure 19 shows the log plot of the accumulated cases and the doubling time calculated using the Excel LINEST function.

Again, we can directly see the positive effect of the Circuit breaker measures that started on April 7, 2020. From a low point of about 5 days the doubling time has increased to about 4 weeks in slightly more than a month.

**Figure 19.**
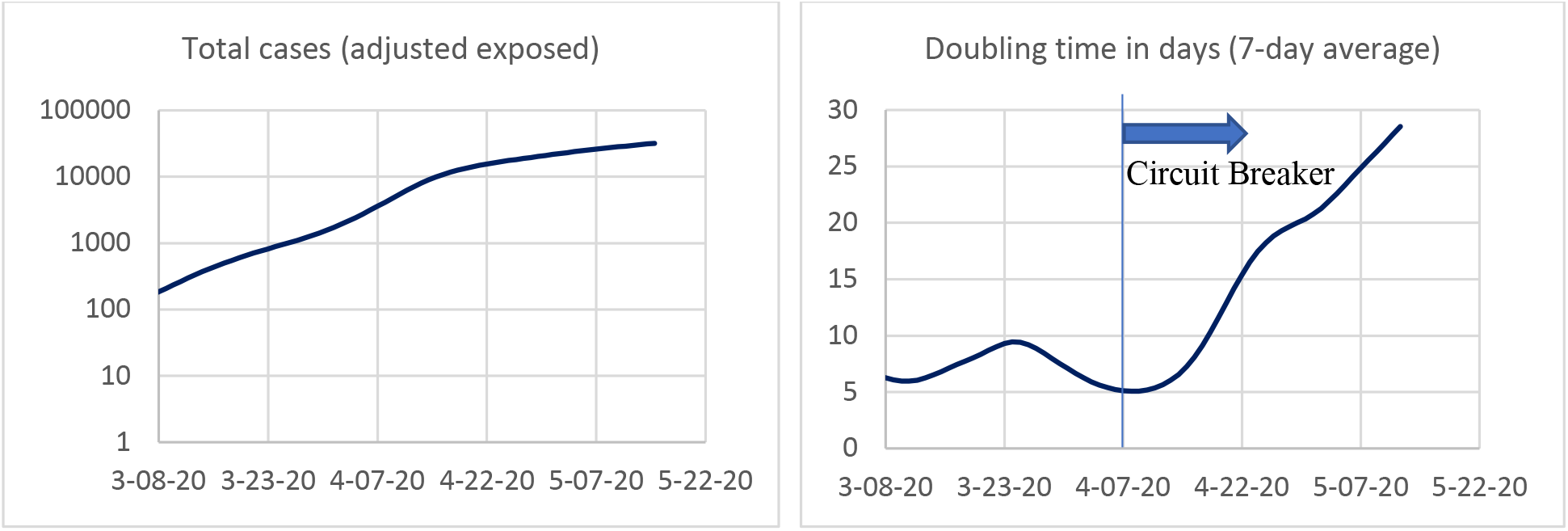
It is easier to see the pronounced effect of the Singapore Circuit breaker on a plot of the doubling time (right) than the accumulated cases plot (left).

## 8. Forecasting the final total number of cases and deaths

When the growth rate is slowing down, we can project the final total cases and death counts by fitting publicly available data to the logistic model. The logistic model is often used to describe the shape of the cumulative epidemic curve (Figure 20) where the number of infected cases grow exponentially at first, then slows down, and finally flattens to a maximum limit. The final epidemic size can be estimated based on this slowing growth.

**Figure 20.**
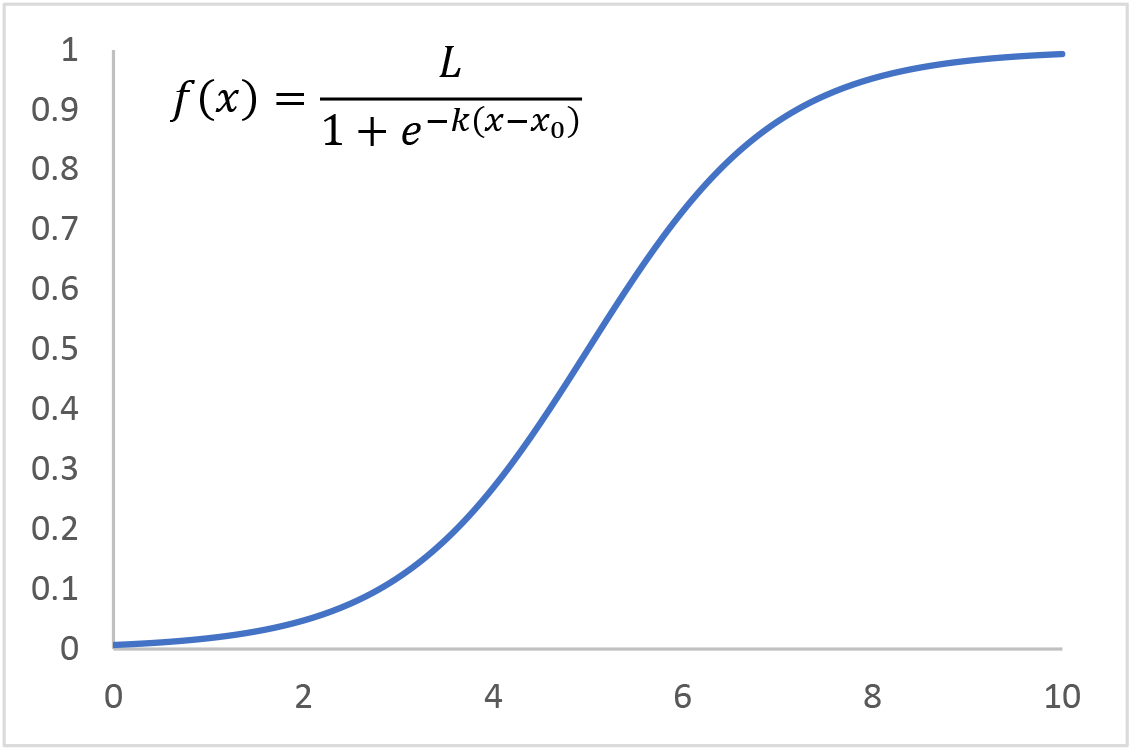
A logistic function. *L* = 1, *k* = 1, *x*_0_ = 5.

For our application, the total number of cases at time *t* can be approximated by ([15])

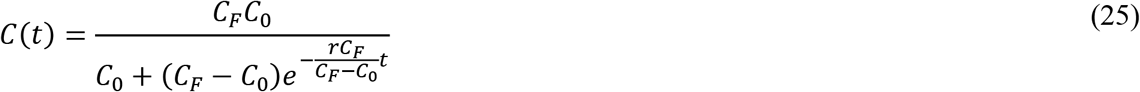

where *r* is the exponential growth rate, *C*_0_ and *C_F_* are the initial and final numbers, respectively. To find the best fit, we use the maximum likelihood method to estimate *C_F_* (see equations (2) and (3)). The parameter is estimated over a relatively short rolling window of, say 14 days, to obtain a moving update. See Figure 21.

**Figure 21.**
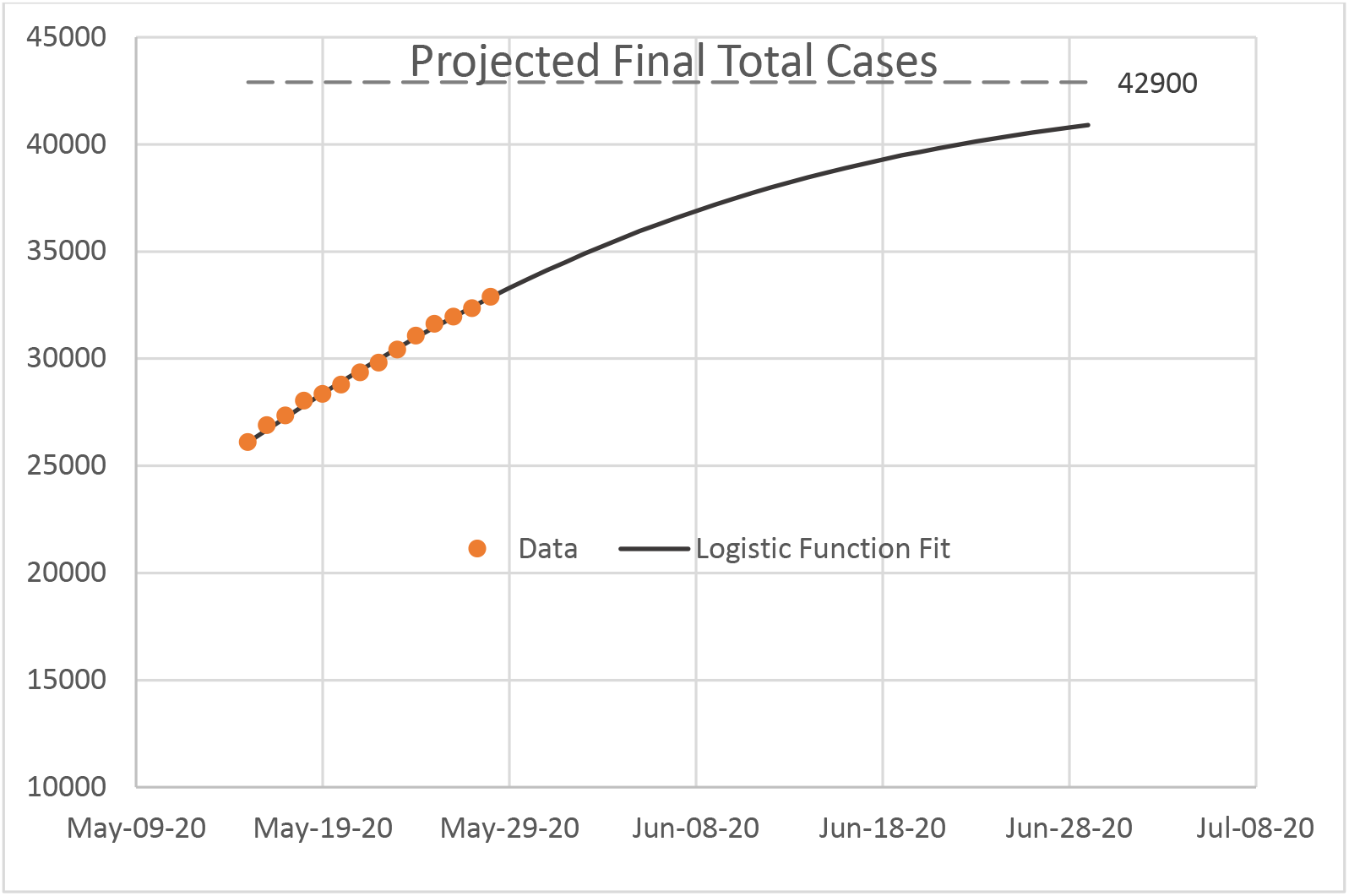
A logistic function is used to fit a two-week data set to project the final total cases.

With an estimate of the final number of cases, we can combine it with the data on case fatality rate to further estimate the final death counts. The case fatality rate varies from region to region but in many countries with a high number of cases they tend to stabilize over time (Figure 22). For example, the case fatality rate for Singapore has remained quite stable at slightly below 1 per 1000 infected cases. Therefore, the final death count is projected to be around 35, assuming a final case number of 43,000. According to the data on deaths per 100 cases in Table 3, Singapore has one of the lowest COVID-19 case fatality rates in the world.

**Figure 22.**
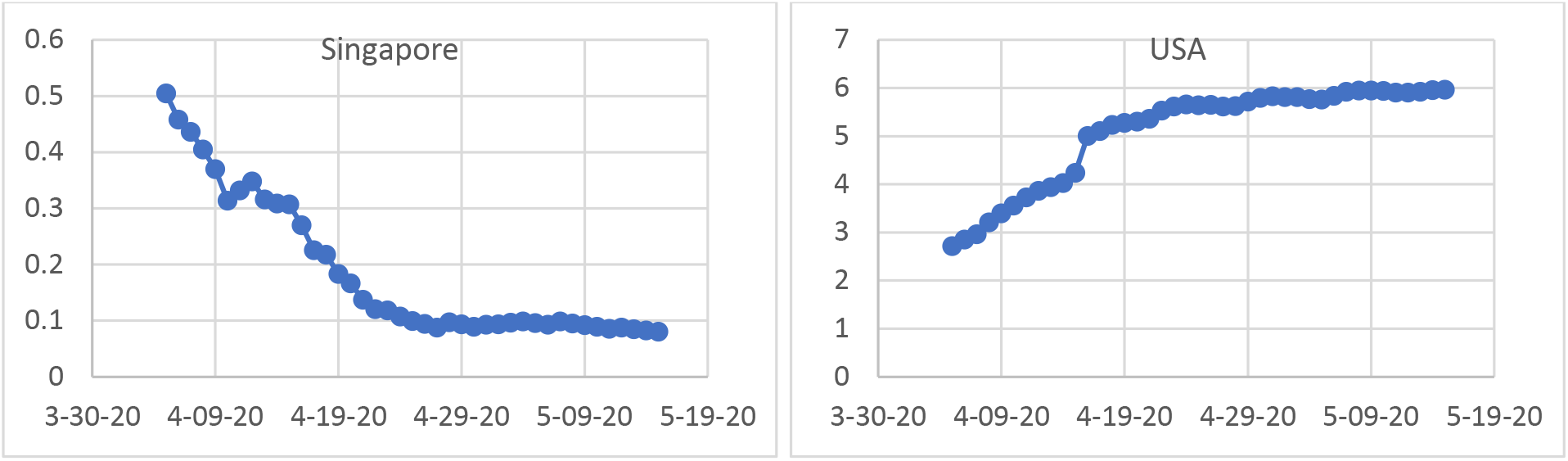
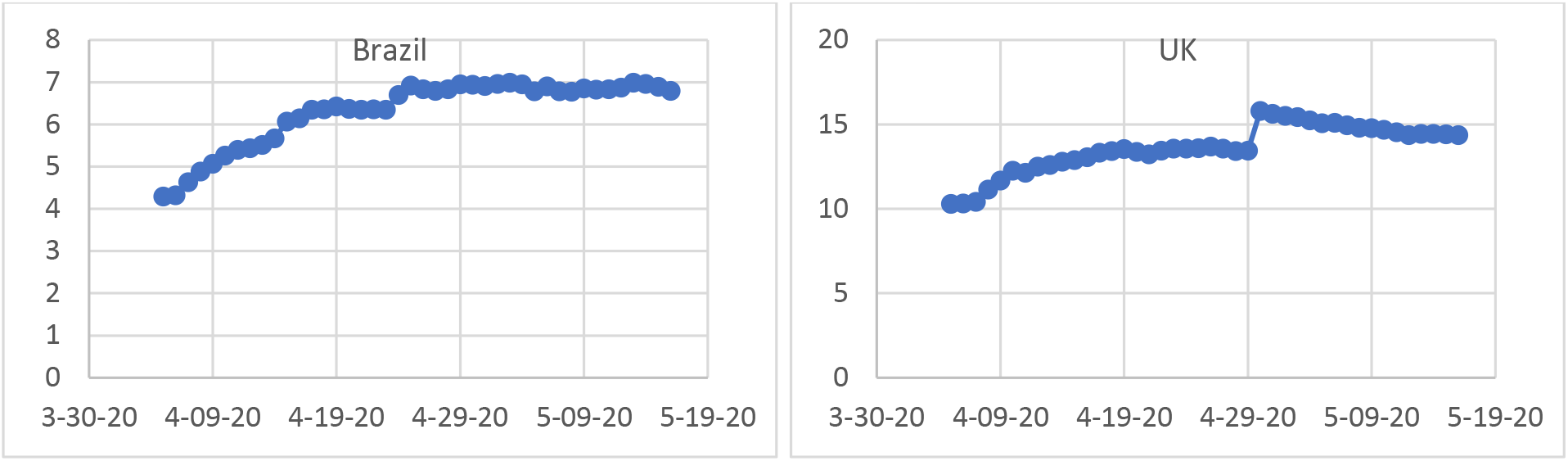
Deaths per 100 cases show a trend toward an equilibrium.

**Table 3.**
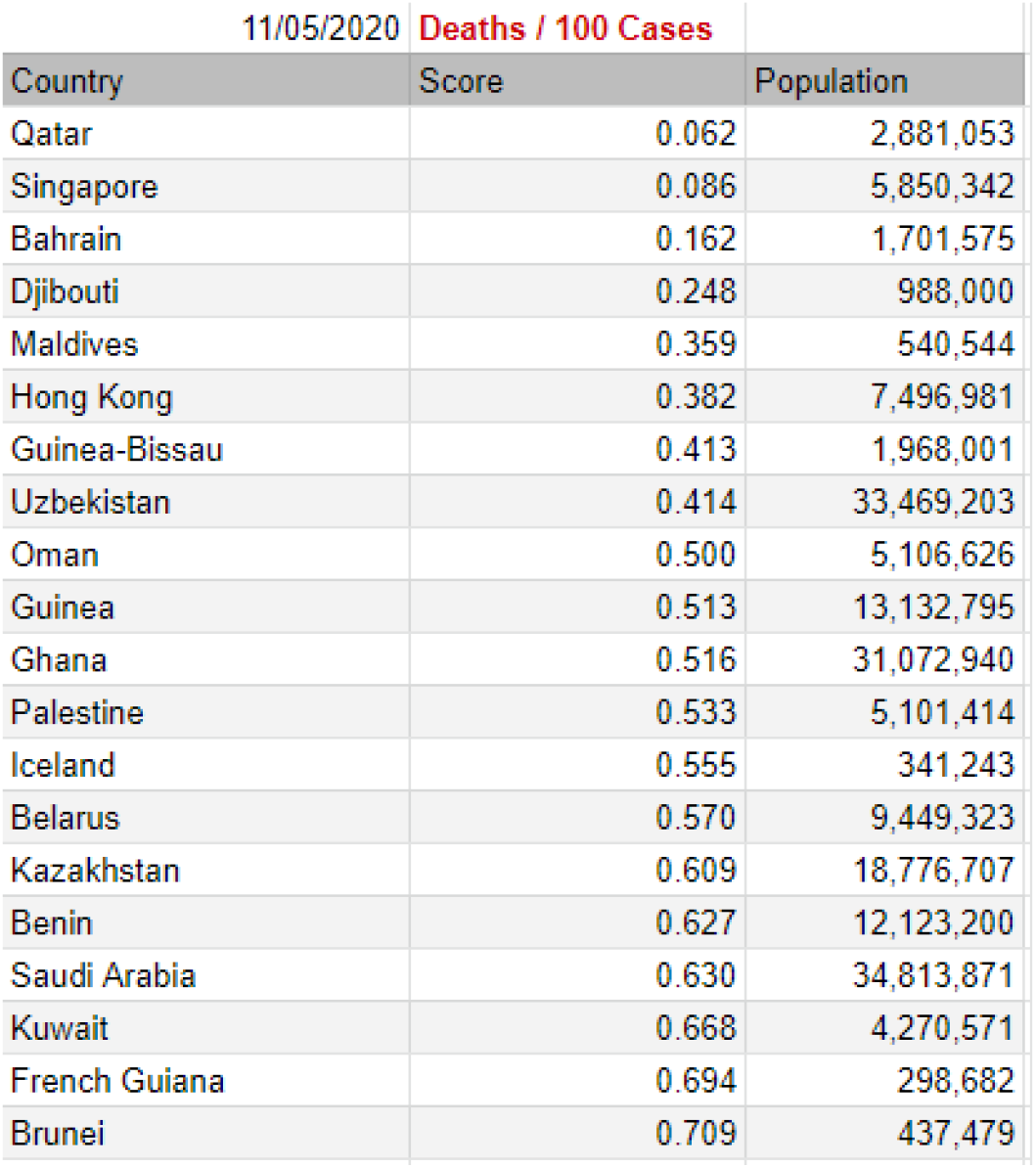
The 20 countries with the lowest case fatality rate.

Table 4 shows the forecasts for a few countries including Singapore and how they compare with the projections by the Institute for Health Metrics and Evaluation (IHME) [26] at University of Washington Medicine. The projections by the IHME are based on more complex analytics and take into account factors such as changes in social distancing measures, diagnostic capability, and hospital capacity. Given that we did not directly account for these factors, our forecasts of the total number of cases and deaths may be considered indicative only. On the other hand, our estimation of reproduction numbers and doubling times based on the current data can be considered reliable and valid. Assuming current prevailing conditions in the populations, results from the COVID-19 Epidemic Calculator are likely to be realistic estimates.

**Table 4.** A comparison of the projected total deaths from the COVID-19 calculator and the Institute for Health Metrics and Evaluation (IHME).

| Country | $C_F$ | Case fatality rate | Projected total deaths using COVID-19 calculator | IHME projection (as at 5-30-20) |
| --- | --- | --- | --- | --- |
| Singapore | 43,000 | 0.08% | 35 | N/A |
| USA | 2,255,800 | 5.9% | 133,000 | 135,109<br>(123,344-157,715) |
| UK | 340,900 | 14.0% | 47,700 | 44,389<br>(42,284-47,610) |
| Brazil | 782,700 | 6.1% | 47,700 | 125,833<br>(68,311-221,078) |

## 9. Conclusions

This paper describes the methods underlying the online COVID-19 Epidemic Calculator for tracking COVID-19 growth parameters. From publicly available data, the calculator is used to estimate the distributions at time of symptom-onset and infection, effective reproduction number, doubling time, final epidemic size, and death toll for Singapore and other countries.

The calculator and the associated graphs clearly show that the Circuit breaker measures imposed from April 7, 2020 in Singapore had an immediate effect of rapidly slowing down the spread of the COVID-19. Additionally, the results also reveal that the effective reproduction number has settled to around 1.0 after about two weeks. Since then, it has remained at that level. This indicates that the infection rate among the dormitory residents is sustained and not likely to be reduced until this group become less susceptible.

The COVID-19 Epidemic Calculator is available in the form of an online Google Sheet [30] that imports daily infection data from the European Centre for Disease Prevention and Control [31]. The results are presented online as dashboards on Tableau Public [32] (Figure 23). It has the advantage of fast execution time without the need for any specialized software package or programming script. Users can also interact with the models by changing the parameters. Comparing with other similar work, our parameter estimates are found to be in good agreement with those estimated using different models and software. By making the COVID-19 Epidemic Calculator readily accessible online, it is hoped that the public and interested learners have the tool to meaningfully assess our effort in fighting COVID-19.

**Figure 23.**
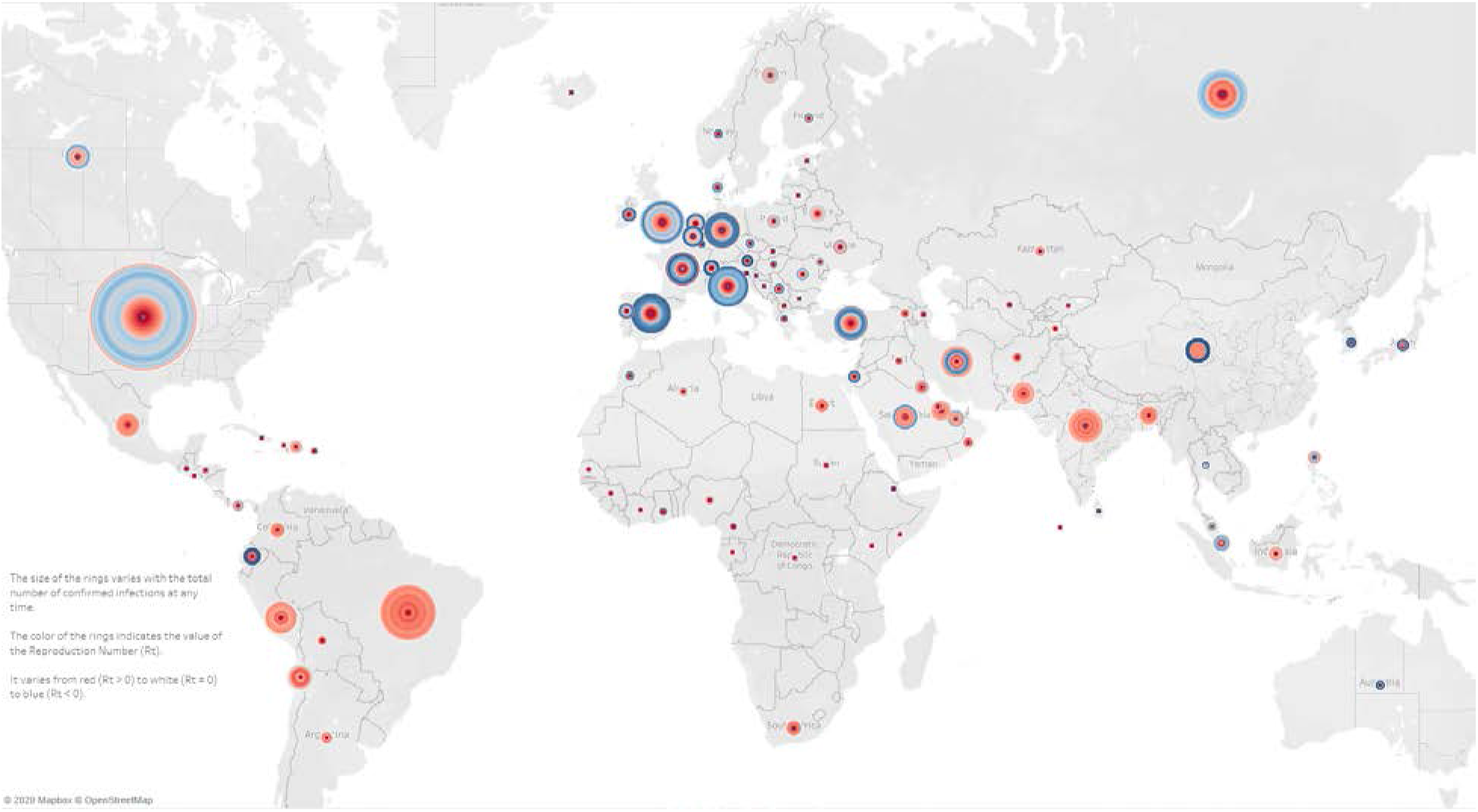
A visualization of the effective reproduction number for countries on a map. The size of each circle is proportional to the total number of infections. The color of the rings within a circle varies over time from red (*R* > 0) to white (*R* = 0) to blue (*R* < 0), reflecting the rate of growth of the virus.

## Data Availability

The data used in the paper are publicly available and cited in the list of references.

http://www.cv19.one

## Author contributions

Conceptualization: Fook Fah YAP and Minglee YONG

Methodology: Fook Fah YAP and Minglee YONG;

Formal analysis and investigation: Fook Fah YAP;

Writing - original draft preparation: Fook Fah YAP;

Writing - review and editing: Fook Fah YAP and Minglee YONG

## Notes

### Competing Interest Statement

The authors have declared no competing interest.

### Funding Statement

No external funding was received.

### Author Declarations

The paper is focused on modeling of publicly available epidemiological data about COVID-19 infections in humans around the world.

## References

1. COVID-19 Situation Report, Ministry of Health Singapore [Internet]. Available from https://covidsitrep.moh.gov.sg/

2. Our World in Data. Coronavirus Source Data [Internet]. https://ourworldindata.org/coronavirus-source-data

3. The COVID Tracking Project [Internet] https://covidtracking.com/api

4. Lauer, Stephen A., et al. “The incubation period of coronavirus disease 2019 (COVID-19) from publicly reported confirmed cases: estimation and application.” Annals of internal medicine (2020).

5. Pung, Rachael, et al. “Investigation of three clusters of COVID-19 in Singapore: implications for surveillance and response measures.” The Lancet (2020).

6. Li, Qun, et al. “Early transmission dynamics in Wuhan, China, of novel coronavirus-infected pneumonia.” New England Journal of Medicine (2020).

7. Nishiura, Hiroshi, Natalie M. Linton, and Andrei R. Akhmetzhanov. “Serial interval of novel coronavirus (COVID-19) infections.” International journal of infectious diseases (2020).

8. Du, Z., et al. “The serial interval of COVID-19 from publicly reported confirmed cases. medRxiv.” DOI: https://doi.org/10.1101/2020.0219 (2020).

9. Vink, Margaretha Annelie, Martinus Christoffel Jozef Bootsma, and Jacco Wallinga. “Serial intervals of respiratory infectious diseases: a systematic review and analysis.” American journal of epidemiology 180.9 (2014): 865–875.

10. World Health Organization. Report of the WHO-China Joint Mission on Coronavirus Disease 2019 (COVID-19) 16-24 February 2020 [Internet]. Geneva: World Health Organization; 2020 Available from: https://www.who.int/docs/default-source/coronaviruse/who-china-joint-mission-on-covid-19-finalreport.pdf

11. Wei WE, Li Z, Chiew CJ, Yong SE, et al. Presymptomatic Transmission of SARS-CoV-2 — Singapore, January 23-March 16, 2020. MMWR, 1 April 2020/69

12. Wu, Joseph T., Kathy Leung, and Gabriel M. Leung. “Nowcasting and forecasting the potential domestic and international spread of the 2019-nCoV outbreak originating in Wuhan, China: a modelling study.” The Lancet 395.10225 (2020): 689–697.

13. Shi, Pengpeng, Shengli Cao, and Peihua Feng. “SEIR Transmission dynamics model of 2019 nCoV coronavirus with considering the weak infectious ability and changes in latency duration.” medRxiv (2020).

14. Chen, Yi-Cheng, et al. “A Time-dependent SIR model for COVID-19 with Undetectable Infected Persons.” arXivpreprint arXiv:2003.00122 (2020).

15. Ma, Junling. “Estimating Epidemic Exponential Growth Rate And Basic Reproduction Number.” Infectious Disease Modelling (2020).

16. Abadie, Alberto, Paolo Bertolotti, and Ben Deaner Arnab. “Epidemic Modeling and Estimation.”

17. Wallinga, Jacco, and Marc Lipsitch. “How generation intervals shape the relationship between growth rates and reproductive numbers.” Proceedings of the Royal Society B: Biological Sciences 274.1609 (2007): 599–604.

18. Wallinga, Jacco, and Peter Teunis. “Different epidemic curves for severe acute respiratory syndrome reveal similar impacts of control measures.” American Journal of epidemiology 160.6 (2004): 509–516.

19. Feller, William. An introduction to probability theory and its applications. Vol. 1. John Wiley & Sons,, 1968.

20. Butler, Ken, and Michael A. Stephens. “The distribution of a sum of independent binomial random variables.” Methodology and Computing in Applied Probability 19.2 (2017): 557–571.

21. Kevin Systrom. “The Metric We Need to Manage COVID-19. Rt: the effective reproduction number” [Internet]. http://systrom.com/blog/the-metric-we-need-to-manage-covid-19/

22. Bettencourt, Luis MA, and Ruy M. Ribeiro. “Real time bayesian estimation of the epidemic potential of emerging infectious diseases.” PLoS One 3.5 (2008).

23. Cori, Anne, et al. “A new framework and software to estimate time-varying reproduction numbers during epidemics.” American journal of epidemiology 178.9 (2013): 1505–1512.

24. Epiforecasts [Internet]. https://epiforecasts.io/covid/

25. Muniz-Rodriguez, Kamalich, et al. “Epidemic doubling time of the COVID-19 epidemic by Chinese province.” medRxiv (2020).

26. Institute for Health Metrics and Evaluation (IHME) COVID-19 Projections [Internet]. https://covid19.healthdata.org/

27. Yang, Zifeng, et al. “Modified SEIR and AI prediction of the epidemics trend of COVID-19 in China under public health interventions.” Journal of Thoracic Disease 12.3 (2020): 165.

28. Dietz, Klaus. “The estimation of the basic reproduction number for infectious diseases.” Statistical methods in medical research 2.1 (1993): 23–41.

29. Bi, Qifang, et al. “Epidemiology and transmission of COVID-19 in 391 cases and 1286 of their close contacts in Shenzhen, China: a retrospective cohort study.” The Lancet Infectious Diseases (2020).

30. COVID-19 Epidemic Calculator [Internet]. https://docs.google.com/spreadsheets/d/1e6QW3QNwLJ3aS2zn_YaYg3N_2408F36dEJ2n_SpwKB8/edit?usp=sharing

31. Data on the geographic distribution of COVID-19 cases worldwide [Internet]. https://www.ecdc.europa.eu/en/publications-data/download-todays-data-geographic-distribution-covid-19-cases-worldwide

32. Global Covid19 Reproduction Number and Doubling Time Tracker [Internet]. http://www.cv19.one

33. Boelle, Pierre-Yves, and Thomas Obadia. “R0: estimation of R0 and real-time reproduction number from epidemics.” Rpackage version 1.2 6 (2015). [Internet]. https://cran.r-project.org/web/packages/R0/index.html

